# Genetic risk factors have a substantial impact on healthy life years

**DOI:** 10.1101/2022.01.25.22269831

**Authors:** Sakari Jukarainen, Tuomo Kiiskinen, Aki S. Havulinna, Juha Karjalainen, Mattia Cordioli, Joel T. Rämö, Nina Mars, FinnGen, Kaitlin E. Samocha, Hanna M. Ollila, Matti Pirinen, Andrea Ganna

**Affiliations:** Institute for Molecular Medicine Finland (FIMM), University of Helsinki, Helsinki, Finland; Finnish Institute for Health and Welfare, Helsinki, Finland; Program in Medical and Population Genetics, Broad Institute of MIT and Harvard, Cambridge, MA, USA; Analytic and Translational Genetics Unit, Massachusetts General Hospital, Boston, MA, USA; Center for Genomic Medicine, Massachusetts General Hospital, Boston, MA, USA; Anesthesia, Critical Care, and Pain Medicine, Massachusetts General Hospital and Harvard Medical School, Boston, MA, USA; Department of Public Health, University of Helsinki, Helsinki, Finland; Helsinki Institute for Information Technology HIIT and Department of Mathematics and Statistics, University of Helsinki, Helsinki, Finland

## Abstract

The impact of genetic variation on overall disease burden has not been comprehensively evaluated. Here we introduce an approach to estimate the effect of different types of genetic risk factors on disease burden quantified through disability-adjusted life years (DALYs, “lost healthy life years”). We use genetic information from 735,748 individuals with registry-based follow-up of up to 48 years. At the individual level, rare variants had higher effects on DALYs than common variants, while common variants were more relevant for population-level disease burden. Among common variants, rs3798220 (*LPA*) had the strongest effect, with 1.18 DALYs attributable to carrying 1 vs 0 copies of the minor allele. Belonging to top 10% vs bottom 90% of a polygenic score for multisite chronic pain had an effect of 3.63 DALYs. Carrying a deleterious rare variant in *LDLR*, *MYBPC3*, or *BRCA1*/*2* had an effect of around 4.1-13.1 DALYs. The population-level disease burden attributable to some common variants is comparable to the burden from modifiable risk factors such as high sodium intake and low physical activity. Genetic risk factors can explain a sizeable number of healthy life years lost both at the individual and population level, highlighting the importance of incorporating genetic information into public health efforts.

Results of the study can be explored at: https://dsge-lab.shinyapps.io/daly_genetics/

## Introduction

Genome-wide association studies (GWAS) have identified thousands of variants associated with biological traits and diseases^1^. Overall, these results demonstrate widespread pleiotropy (i.e., genetic variants associated with more than one trait)^2^. Studies commonly quantify the impact of genetic variation on a single disease at a time^3–5^, or when considering multiple diseases^6–8^ do not use a single metric that can capture overall disease burden. It is therefore challenging to assess the impact of genetic variation on overall health and to compare the total impact of different variants.

Past efforts in comparative risk assessment involve quantifying the effects of modifiable adverse exposures (e.g., sodium intake) on health outcomes to inform public health measures^9^. For genetic risk factors this type of assessment has not been systematically performed, perhaps owing to lack of interventions. However, advances in human genetics (e.g., polygenic scores, PGS) have generated increasing interest in using an individual’s genetic risk in prioritization of screening (e.g., for cancers) and primary prevention (e.g., for coronary artery disease)^7, 10–12^, making information on genetic risk clinically actionable. Also, genetic risk factors are becoming modifiable as PGSes have been controversially evaluated for their potential in embryo selection^13–15^ and *in vivo* gene editing is progressing towards clinical application^16–18^. Thus, there is a need for a comparative risk assessment framework of genetic risk factors.

One prominent metric for disease burden is the disability-adjusted life year (DALY). DALYs represent the loss of healthy life years through worsened quality of life and premature death attributable to a disease^19^. Combining both quality of life and mortality into a single metric, DALYs are used to monitor disease burden across hundreds of countries in the Global Burden of Disease study (GBD)^9, 19^. The GBD estimates the yearly amount of DALYs in each country attributable to a list of collectively exhaustive and mutually exclusive diseases and injuries^19^. DALYs are the sum of years lived with disability (YLDs, “lowered quality of life”) and years of life lost (YLLs, “premature death”) (**Extended Data Fig. 1**). Simplifying, the yearly YLDs are estimated by multiplying the prevalence of a disease by its disability weight, which represents the magnitude of health loss due to living with the disease scaled between 0 (perfect health) and 1 (death). Yearly YLLs are estimated by multiplying the number of deaths attributable to a disease by the standard life expectancy at age of death^19^.

We propose a new approach for combining genetic association results for 80 diseases from two biobank studies with DALY estimates from the 2019 GBD study^19^ to provide an overview of the impact of genetic variation on lost healthy life years both at individual and population level. We rank different genetic risk factors in terms of their total health impact and compare genetic risk factors with traditional modifiable risk factors, presenting a template for comparative risk assessment of genetic risk factors.

## Results

### Estimating attributable DALYs

Our method is similar to the GBD approach which estimates the disease burden attributable to *modifiable* risk factors/exposures^9^ except here we consider different classes of *genetic* risk factors: common variants, rare deleterious variants, HLA alleles, APOE haplotypes, and PGSes (**Fig. 1**), referred to as *genetic exposures*. An advantage over the GBD risk factors approach^9^ is that our genetic association results are estimated using individual-level data from two large population-based biobank studies: FinnGen (n=309,136) and UK Biobank (UKB, n=426,612) with registry-based follow-up of 48.7 and 22.4 years respectively. Also, with some important caveats, the estimates we present allow for a causal interpretation by virtue of genetic exposures having fewer possible confounders than modifiable risk factors. In total we considered 80 noncommunicable diseases that account for 83.1% of the total DALYs out of all noncommunicable diseases in Finland 2019^19^ see **Supplementary** Tables 2 and 3 (**ST2-ST3**).

**Fig. 1:**
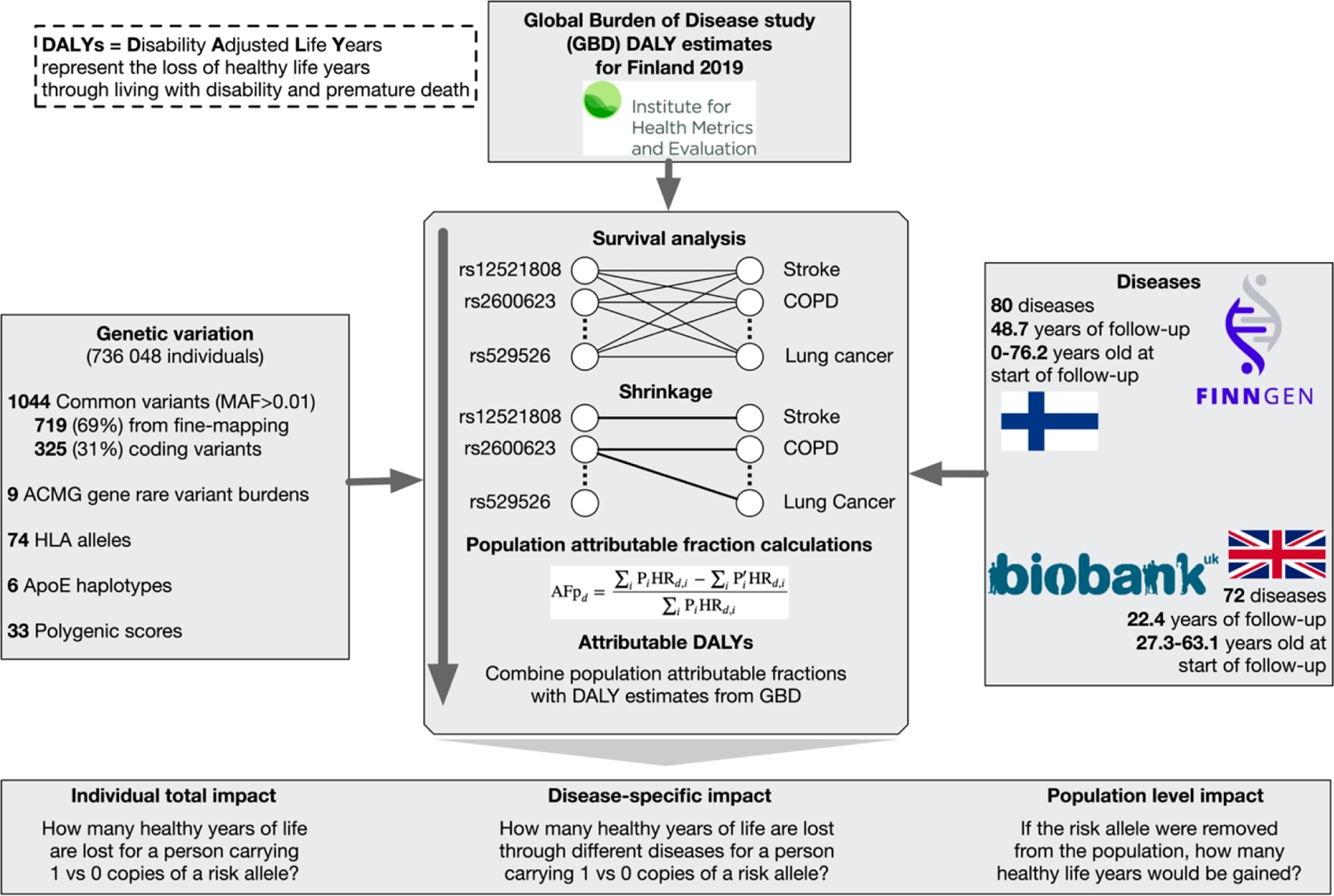
Study overview. GBD, Global Burden of Disease study; MAF, minor allele frequency; HLA, human leukocyte antigen system; ACMG, American College of Medical Genetics; AFp, population attributable fraction; HR, hazard ratio.

For each genetic exposure-disease pair, we estimated the hazard ratio (HR) using a Cox proportional hazards model. Because a single genetic variant is expected to be associated with only a minority of the considered 80 diseases, we used a shrinkage approach with a spike-and-slab type prior distribution for the log-HRs of each genetic exposure for the 80 diseases (**Methods**). We discarded any genetic exposure-disease associations where the posterior probability of the null model was above 10%.

Overall, we estimated the HRs through 92,800 survival analyses (associations of 1044 common variants, 9 rare variant burdens, 74 HLA alleles, and 30 PGSes with 80 diseases) and, after shrinkage, retained 3,123 HRs for genetic exposure-disease pairs, most of which (67.1%) were genome-wide significant (*P*<5×10^-8^) and 99% had an association with *P*<7.3×10^-4^ (**Extended Data Fig. 2**). Using the HR estimates and frequencies of the genetic exposures, we estimated the population attributable fraction of disease cases for each genetic exposure (the proportion of disease cases that would be prevented if the exposure was removed). We combined these fractions with disease-specific population DALYs for Finland 2019 from the GBD^19^ (**ST3**) to obtain attributable DALY estimates (**Methods**). Finally, we summed attributable DALYs across the 80 diseases to estimate the total impact of genetic exposures. The total individual attributable DALYs, our main measure of interest, can be interpreted as the *expected loss of healthy life years for an individual attributable to having a certain genetic exposure at birth*. Because we consider different types of genetic exposures the exact definition of attributable DALYs varies by exposure type. Because the disease definitions in GBD are not overlapping and the DALYs are comorbidity-corrected^19^, the final estimates take double-counting into account.

### Attributable DALYs for common variants

For all variants, we defined the minor allele as the less common allele in FinnGen. We considered 1,044 independent common variants (minor allele frequency, MAF > 1%). We selected 564 of these based on having at least one *P*<5×10^-8^ association with any of the 80 diseases and having the highest probability of being causal within a SuSiE fine-mapped^20^ 95% credible set in FinnGen. Additionally, we selected 155 common variants with at least one *P*<5×10^-12^ association with 6 traditional risk factor traits (BMI, HbA1c, HDL cholesterol, LDL cholesterol, systolic blood pressure, cigarettes per day) that had the highest probability of being causal within a within a SuSiE fine-mapped 95% credible set in UKB^21^. Last, we included 325 coding variants having a *P*<5×10^-8^ association with one of the diseases in FinnGen. Among the 1044 variants, 34.6% were annotated as missense (n=335) or putative loss-of-function (n=26, pLOF). The HRs for common variants were comparable between FinnGen and UKB (**Extended Data Fig. 3**) and we consequently meta-analyzed the results. All estimates are for the comparison of 1 vs 0 copies of the minor allele, so the individual attributable DALYs thus correspond to the *expected loss of healthy life years if an individual with 0 copies of the minor allele had instead 1 copy at birth*.

Overall, carrying 1 vs 0 copies of the common variants we studied resulted in relatively small effects on lost healthy life years in terms of DALYs, with only 56 out of 1,044 (5.4%) variants having over 0.25 attributable DALYs (**ST11-ST12**). Many of the top hits for attributable DALYs were in chromosome 6, both inside and outside of the HLA region (**Fig. 2a**). We imputed the 7 classical HLA genes at the two-field resolution level (i.e., unique protein sequence level) using a Finnish-specific reference panel^22^ and provide attributable DALYs for HLA alleles in (**Extended Data Fig. 4, ST9**). However, we caution about the interpretation of attributable DALYs in this context because, for multiallelic loci, the estimates are not straightforward to interpret.

**Fig. 2:**
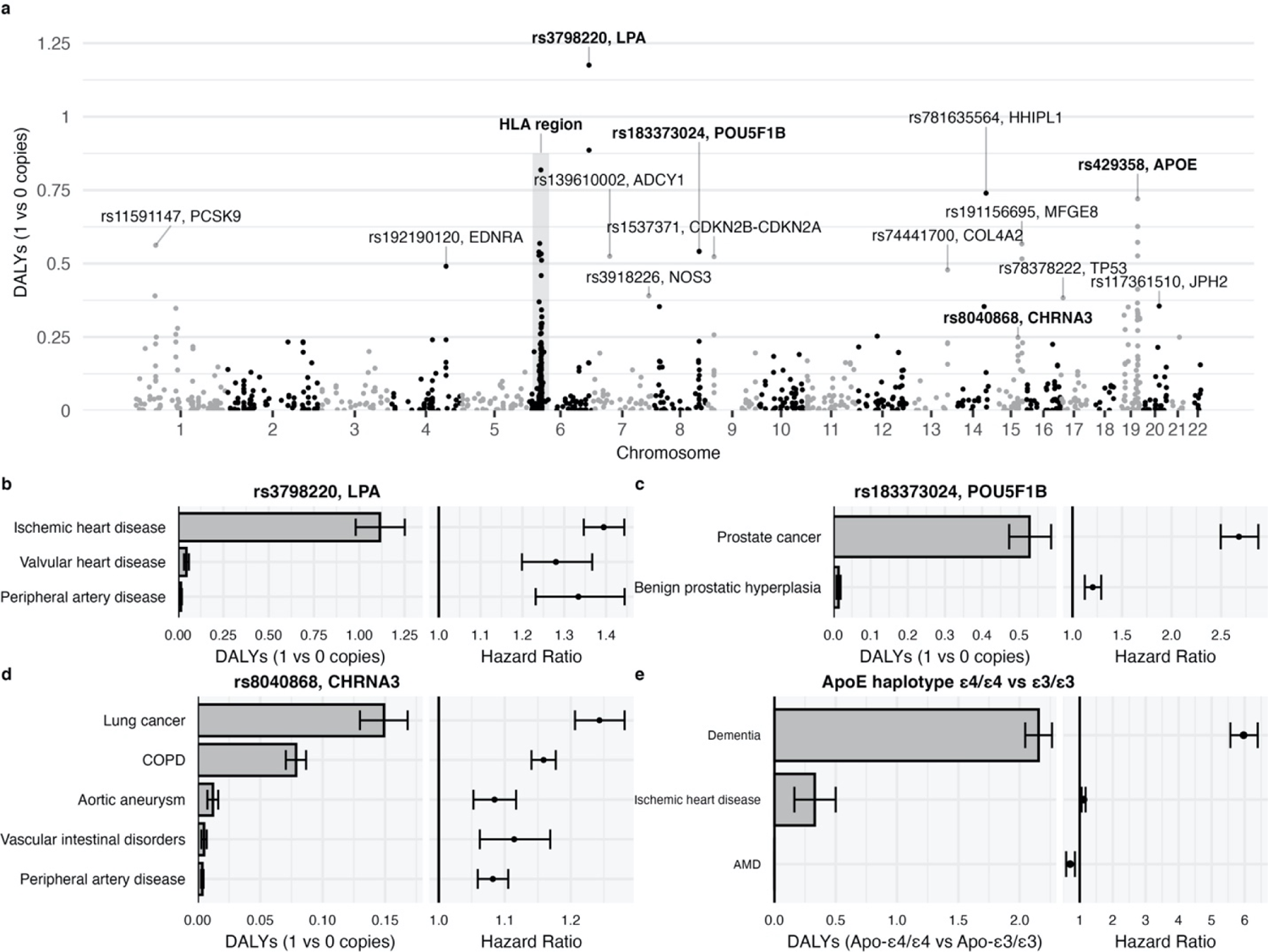
Effect of common variants on DALYs. a, Absolute effect on DALYs from carrying 1 vs 0 copies of the minor allele for each common variant. We analyzed separately imputed alleles in the HLA region. Results for this region are provided in (Extended Data Fig. 4). b-e, For 3 common variants and the APOE haplotypes (in bold in panel a) we reported attributable DALYs and HRs separately for each disease. Error bars denote 95% confidence intervals. COPD, chronic obstructive pulmonary disease; AMD, age-related macular degeneration.

The variant with the highest number of attributable DALYs was rs3798220, a missense variant in the *LPA* gene, with 1.18 (95% confidence interval (CI) 1.03-1.32, *P*=1.3×10^-58^) attributable DALYs from carrying 1 vs 0 copies of the C allele (**Fig. 2B**). The effect was almost exclusively through ischemic heart disease (1.11 DALYs) and to a lesser extent through non-rheumatic valvular heart disease (0.046 DALYs) and lower extremity peripheral artery disease (0.016 DALYs) despite similar risk increases. This is because of the larger number of population DALYs attributed to ischemic heart disease by the GBD (e.g., 60-fold difference to lower extremity peripheral artery disease, **ST3**), illustrating the importance of using measures other than just relative risk.

One interesting example is rs183373024, a non-coding variant near the *POU5F1B* gene^23^, with 0.54 (95% CI 0.48-0.60, *P*=1.1×10^-74^) attributable DALYs mainly through prostate cancer (**Fig. 2c**). Another example is rs8040868, a synonymous variant in the well-known *CHRNA5/A3/B4* gene cluster associated with nicotine dependence^24^, with 0.25 (95% CI 0.23-0.27, *P*=1.6×10^-95^) attributable DALYs (**Fig. 2d**), with effects through lung cancer, COPD, aortic aneurysm, vascular intestinal disorders, and lower extremity peripheral artery disease (all consequences of smoking).

Given the strong associations between APOE alleles and longevity^25^, we defined the three main APOE alleles determined by two SNPs rs429358 and rs7412. Carrying the most deleterious Apo-ε4/ε4 haplotype instead of the most common Apo-ε3/ε3 resulted in 2.48 (95% CI 2.28-2.68, *P*=1.0×10^-128^) attributable DALYs, mainly through increase in risk of Alzheimer’s disease and other dementias (HR=5.97, 95% CI 5.57-6.40, **Figure 2e**, **ST8**). Overall, out of the top 10% common variants with the highest number of attributable DALYs, 49.4% were significantly associated (nominal *P*<0.05) with longevity in the largest GWAS on lifespan^5^ versus 18% in the bottom 10%.

The full results for the common variant analysis can be explored at: https://dsge-lab.shinyapps.io/daly_genetics/

### Attributable DALYs for rare deleterious variants

Rare deleterious coding variants (MAF<0.001) are often clinically relevant because of their large *individual* (not *population*) level effects. However, past studies quantifying the effect of rare variants have relied on highly selected clinical populations due to lack of population-based whole-exome/genome sequencing data. Recent whole-exome sequencing data from UKB (n=174,379) provides a unique opportunity to address this issue.

The American College of Medical Genetics and Genomics (ACMG) recommend reporting incidental findings in clinical exome and genome sequencing for 73 genes^26, 27^. We estimated the attributable DALYs for two types of burdens for these ACMG genes: 1) putative loss-of-function (pLOF) variant burden and 2) “pathogenic” or “likely pathogenic” Clinvar^28^ variant burden (for *BRCA1*/*2* we used “pathogenic” ENIGMA^29^ variants instead). We report results for genes with at least 35 individuals with a positive burden and at least one retained burden-disease association. Taking the pLOF burdens as an example, the estimated individual attributable DALYs correspond to the *expected loss of healthy life years if an individual carrying no loss-of-function variants in the gene would be instead carrying at least one*.

The 5 genes most impactful in terms of DALYs (**Fig. 3**, **ST15-16**) were *LDLR* (ischemic heart disease), *BRCA2* (breast, ovarian, liver, and prostate cancer, COPD), *MYBPC3* (cardiomyopathy and myocarditis), *BRCA1* (breast and ovarian cancer), and *MLH1* (colon and rectum cancer). As an example, individuals carrying one pLOF in *BRCA1* lose on average 4.08 (95% CI 2.74-6.32, *P*=1.4×10^-5^) healthy life years through breast cancer (HR=7.01, 95% CI 4.94-9.94, DALYs=2.11, 95% CI 1.39-3.14) and ovarian cancer (HR=16.2, 95% CI 8.22-31.8, DALYs=1.97, 95% CI 0.95-3.93).

**Fig. 3:**
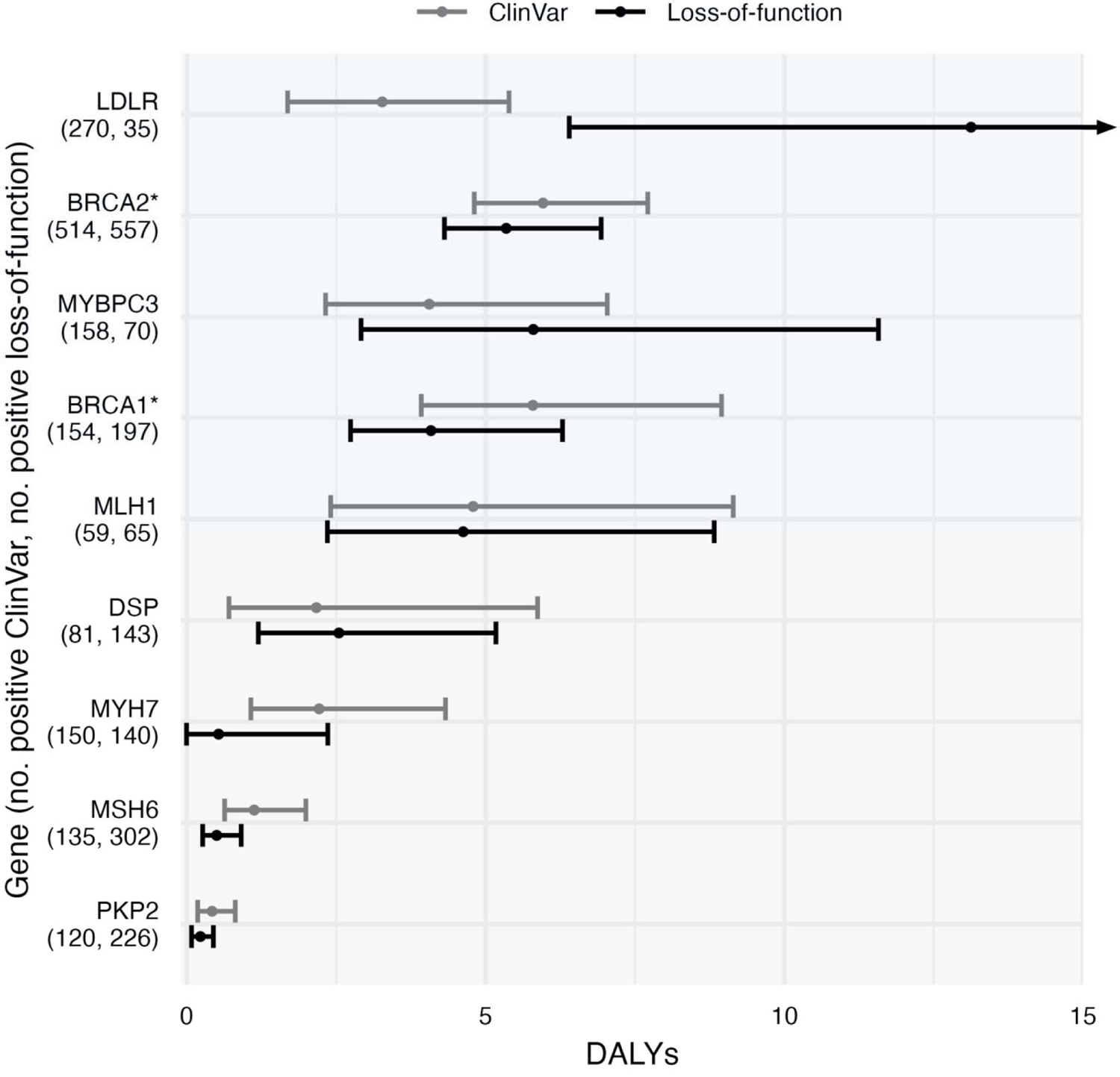
DALYs attributable to carrying a deleterious rare variant in ACMG genes. The ClinVar^28^ burden contains all variants annotated as “pathogenic” or “likely pathogenic”. *For *BRCA1* and *BRCA2* we only considered variants from ENIGMA^29^ annotated as “pathogenic”. The loss-of-function burden contains all variants annotated as putative loss-of-function with high confidence in gnomAD^30^. Error bars denote 95% confidence intervals.

### Attributable DALYs for polygenic scores

We considered 30 PGSes covering major diseases, clinical risk factors, and psychobehavioral traits. We estimated the effect on DALYs attributable to belonging to the top 10% of a PGS versus the bottom 90%, which depict the *expected loss of healthy life years if an individual in the bottom 90% of a PS was instead in the top 10% at birth*. Overall, the attributable DALYs varied from 0.07 (inflammatory bowel disease^31^) to 3.81 (shorter lifespan^32^) (**Fig. 4a**, **ST13-ST14**). Many of the PGSs exhibited significant pleiotropy, with a median of 16 (IQR 9-28) PGS-disease associations remaining after shrinkage. PGSes with the highest number of retained disease associations were multisite chronic pain^33^ (n=44), lower educational attainment^34^ (n=40), and shorter lifespan^32^ (n=40).

**Fig. 4:**
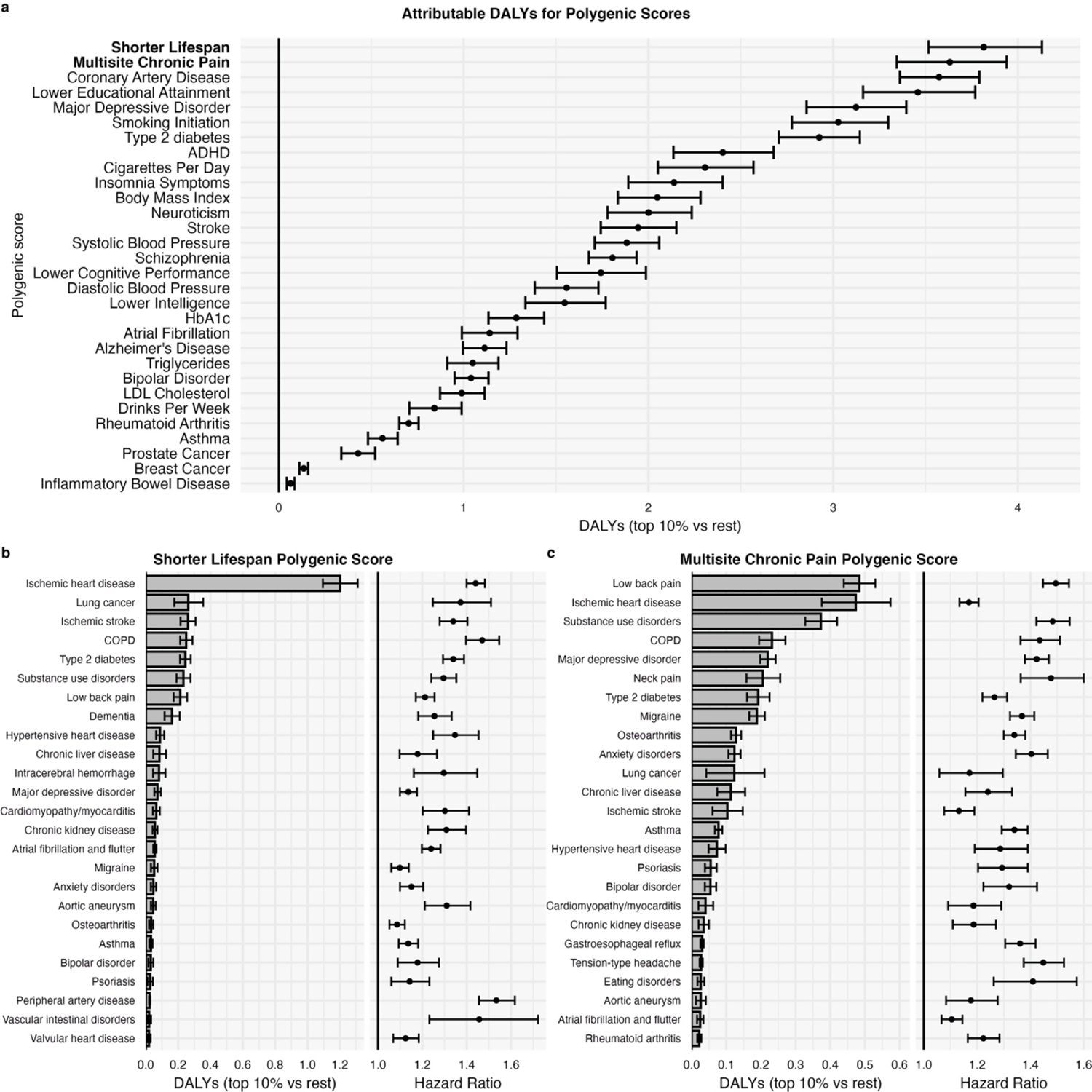
Polygenic score impact on DALYs. **a**, DALYs attributable to belonging in the top 10% vs bottom 90% of each PGS. **b, c**, Top 25 diseases in terms of attributable DALYs and HRs for two PGSes (bold in panel **a**). Error bars denote 95% confidence intervals. ADHD, attention deficit hyperactivity disorder; HbA1c, glycated hemoglobin.

A PGS predicting shorter lifespan^8^ had the highest impact. Individuals in the top 10% of the PGS are expected to lose 3.81 (95% CI 3.52-4.13, *P*=4.6×10^-131^) healthy life years compared to an individual in the bottom 90%. This PGS acts mainly through ischemic heart disease (1.2 DALYs) and to lesser extent through lung cancer, ischemic stroke, COPD, type 2 diabetes, substance use disorders and low back pain, each accounting between 0.21-0.26 DALYs (**Fig. 4b**). Interestingly, a PGS for multisite chronic pain^33^ had the second highest impact at 3.63 (95% CI 3.33-3.93, *P*=5.0×10^-124^) DALYs, mainly through low back pain (0.48 DALYs), ischemic heart disease (0.47), substance use disorder (0.37), COPD (0.23), depression (0.22), and neck pain (0.21) (**Fig. 4c**). Results for the polygenic score analysis can be explored at: https://dsge-lab.shinyapps.io/daly_genetics/

### Sex-specific effects

We repeated all analyses stratified by sex and present estimates for the main analyses (apart from rare variants) by sex in supplements (**ST8-ST9, ST11-ST14**). Significant sex differences in total DALYs at *P*<0.05 were observed for 474 (45%) of the common variants (**Fig. 5a**). Sex differences in attributable DALYs can result from differences in the effect of the genetic exposure on the disease (i.e., differences in HRs) or differences in DALYs attributed to men and women by the GBD^19^. rs738409 (PNPLA3-I148M), a missense variant in *PNPLA3* linked to liver fat accumulation and steatohepatitis^35^, provides a clarifying example: Carrying 1 vs 0 copies of the minor allele resulted in 0.27 (95% CI 0.24-0.30, *P*=1.6×10^-71^) attributable DALYs in males and 0.05 (95% CI 0.03-0.07, *P*=2.5×10^-7^) DALYs in females (sex difference *P*=1.0×10^-34^). The sex difference is in part driven by differences in HR between men and women for chronic liver disease (1.32, 95% CI 1.28-1.37 in males vs 1.21, 95% CI 1.17-1.26 in females, *P* for sex difference = 3.6×10^-4^) and, in part, because DALYs for chronic liver disease are higher in men than women^19^ (431 vs 158 yearly DALYs per 100,000) (**Fig. 5c**).

**Fig. 5:**
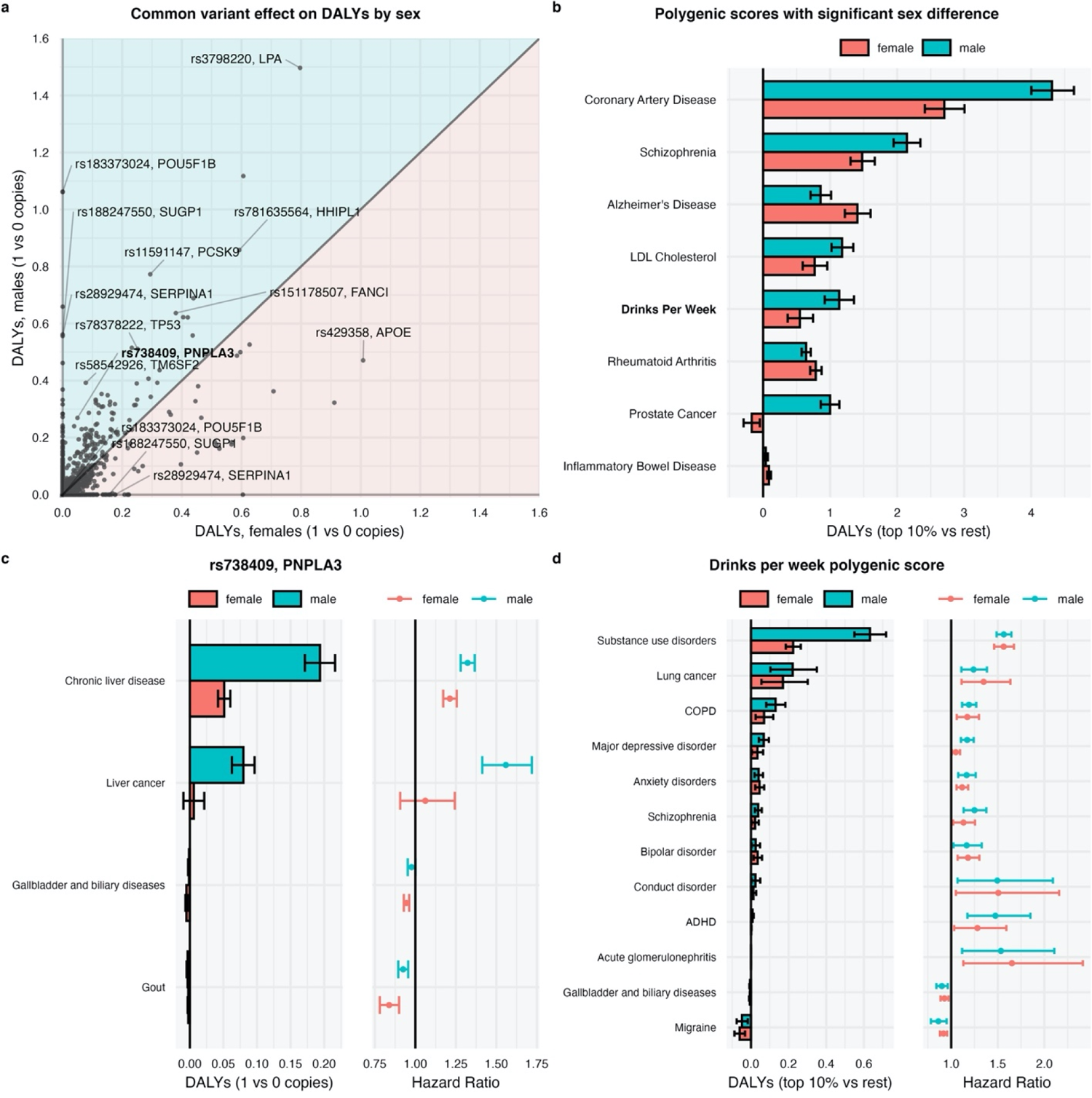
Sex-specific impact of common variants and polygenic scores on DALYs. **a**, Absolute attributable DALYs from carrying 1 vs 0 copies of the minor allele separately for males and females. **b**, Polygenic scores (top 10% vs rest) with a significant sex difference in attributable DALYs. **c**, For rs738409 (*PNPLA3*) (in bold in panel **a**) we report the attributable DALYs and HRs for each disease by sex. **d**, Attributable DALYs and HRs by disease and sex for the PS predicting drinks per week (in bold in panel b). Error bars denote nominal 95% confidence intervals.

8 out of 30 PGSes had significant sex differences in attributable DALYs (**Fig. 5b**). Most of such differences were explained by different DALYs attributed to men and women rather than differences in underlying association estimates. For example, a PGS predicting weekly alcohol consumption had similar HRs between sexes across all diseases (**Fig. 5d**) but markedly different effect on DALYs for substance use disorders reflecting the higher disease burden disease for substance use disorders among men (1,500 yearly DALYS per 100,000) than women (497 yearly DALYs per 100,000) as estimated by the GBD^19^.

### Population attributable DALYs for common variants

So far, our results have been from an individual’s perspective. Next, for the Finnish population, we estimated the amount of attributable *population* DALYs per year per 100,000 from all (heterozygous and homozygous) carriers of the minor allele: *the expected amount of healthy life years per year per 100,000 individuals in the population that would be gained if the minor allele were completely removed*. rs7859727 (*CDKN2B-CDKN2A*) had the highest population attributable DALYs, with minor allele carriers accounting for 447 (95% CI 420-473, *P*=4.1×10^-231^) yearly population DALYs per 100,000 in Finland 2019 (**Fig. 6a**). The large population effect of this variant is explained by its effect on ischemic heart disease (HR=1.17, 95% CI 1.16-1.18) and high frequency in the Finnish population (MAF=41%). Comparing to population DALY estimates for classic risk factors from the GBD^9^ (**Fig. 6a**), the attributable population DALY estimates of several common variants are similar to the total impact of a diet high in sodium (300 yearly population DALYs per 100,000), low physical activity (415), and drug use (595), but substantially less impactful than the most important classic risk factors such as high systolic blood pressure (3666), smoking (2992), and high BMI (2506)^9^.

**Fig. 6:**
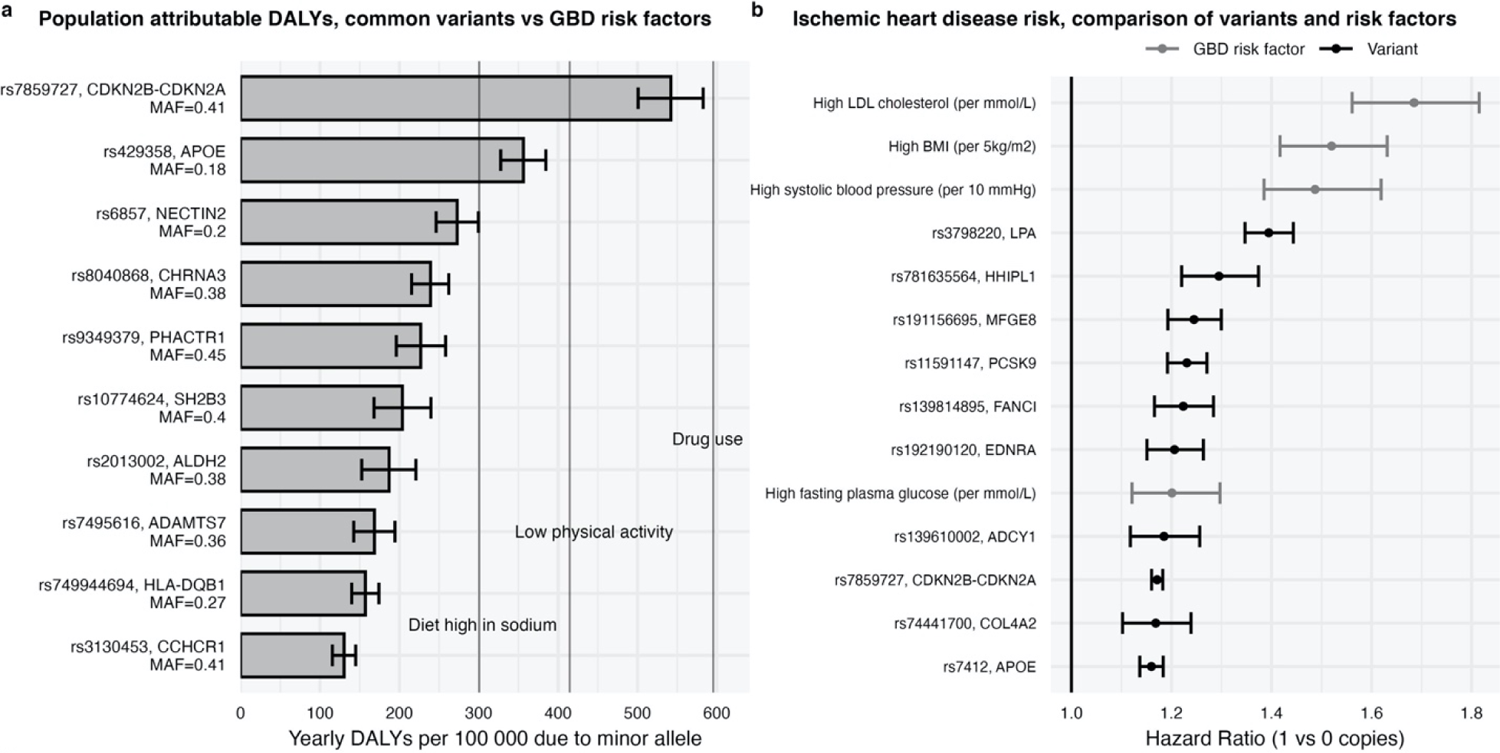
Effect of common variants on population-level DALYs in the Finnish population. a, Bars represent yearly population-level DALYs attributable to top 10 ranking common variants. The vertical lines represent yearly population-level DALYs attributable to three selected classic risk factors as estimated by the GBD^9^ for Finland 2019. b, Top 10 HRs for ischemic heart disease of common variants and 4 HRs of conventional risk factors as estimated by the GBD (50-54 year age group)^9^.

In other words, removing the risk allele rs7859727 from the Finnish population would have a comparable expected impact on increasing healthy life years as changing sodium consumption or physical activity to the theoretical minimum risk level, or completely removing drug use as a problem. To further expand on this concept, we compared the HRs for ischemic heart disease between 8 common variants and 4 clinical risk factors (**Fig. 6b**). Clinically significant changes in traditional risk factors (e.g., 10 mmHg higher systolic blood pressure, 1 mmol/L higher fasting glucose) as estimated by the GBD lead to comparable increases in ischemic heart disease risk (HR=1.20 to 1.69)^9^ as having 1 vs 0 copies of the minor allele for the cardiovascular risk variants (HR=1.16 to 1.39).

### Additional population DALYs attributable to Finnish-enrichment

Finland is a well-known example of an isolated population where multiple historical bottlenecks^36^ have contributed to the enrichment of several functional genetic variants^36, 37^ otherwise rare in non-Finnish populations. It is therefore interesting to estimate the population attributable DALYs that are due to the enrichment in the Finnish population compared to non-Finnish-non-Swedish-non-Estonian European (NFSEE) populations. There were 56 Finnish-enriched variants that had at least 5-fold MAF enrichment in the Finnish ancestry compared to NFSEE ancestry (from gnomAd) where the NFSEE MAF was below 0.01. The largest impact on population attributable DALYs (**Fig. 7**) was observed for rs143473297 (*TOMM40*) contributing to 56.1 (95% CI 50.1-62.0, *P*=6.3×10^−76^) yearly population DALYs per 100,000 individuals of Finnish ancestry through increased risk of dementia (HR=1.95, 0.57 *individual* DALYs, **ST12**). This variant had a 152-fold MAF enrichment in the Finnish population and a negligible effect in NFSEE ancestry. One remarkable example of a protective Finnish-enriched variant is rs191156695, an inframe insertion in *MFGE8*^38^. The presence of the Finnish-enriched allele in the population contributes to *preventing* 39.1 (95% CI 32.0-45.8, *P*=4.6×10^-29^) yearly population DALYs per 100,000 individuals of Finnish ancestry (**Fig. 7**) solely through decreasing ischemic heart disease risk (HR=0.80, 95% CI 0.77-0.84).

**Fig. 7:**
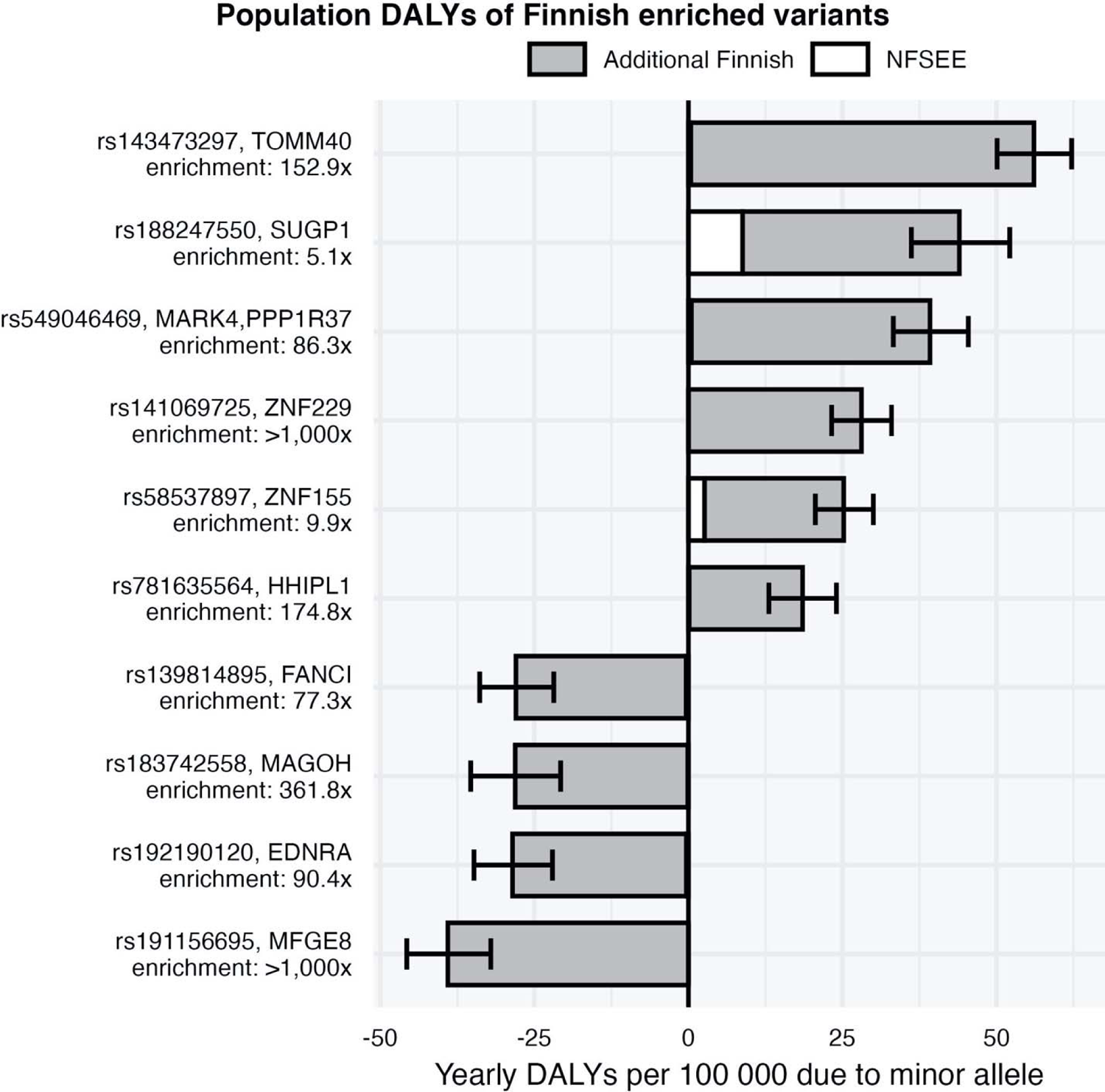
Impact of Finnish-enriched variants on population-level DALYs. Proportion of disease-specific population DALYs accounted for by the 9 Finnish-enriched variants with the largest effects. The white part of the bars denotes the number of population DALYs attributable to the variant if the Finnish MAF were equal to the NFSEE MAF, the grey part is the additional population DALYs resulting from the enrichment in the Finnish population. Error bars denote nominal 95% confidence intervals. NFSEE, non-Finnish-non-Swedish-non-Estonian Europeans. For many variants the contribution in NFSEE is neglectable and white bar not visible.

### Sensitivity analyses

We performed three main sensitivity analyses. First, we explored whether the effect of the genetic exposures on the diseases was age dependent by performing age-stratified survival analyses. Perhaps unsurprisingly^39^, we observed age-varying HRs for genetic exposures (**Extended Data Fig. 5**). For example, for a coronary artery disease PGS^40^, the HRs were 2.50 (95% CI 2.31-2.70) for the 50-54 age group and 1.75 (95% CI 1.62-1.87) for the 70-74 age group.

Second, we examined if accounting for relatedness would result in different estimates for a subset of the survival analyses. For 2562 common variants-disease pairs we estimated the HRs using a survival model clustered by family indicator to generate robust standard errors. Compared to the main analysis estimates, robust standard errors were median 1.0128 (IQR: 1.0044–1.0205) times larger. Thus, accounting for relatedness would not meaningfully affect the confidence intervals and *P*-values.

Third, we examined the performance of our shrinkage approach, showing that it is robust to the choice of different prior parameters (**Extended Data Fig. 6**) and can identify true causal variants in simulated GWAS data (**Extended Data Fig. 7, ST7**). We further compare our shrinkage approach with a conservative colocalization-based approach for 48 of the diseases (**Supplementary Information**). The colocalization approach identified 94 common variants for which two or more diseases co-localize. 82.0% of these variants and the corresponding colocalizing diseases were identified by our shrinkage approach. Conversely, only 22.4% of pairs of diseases reported by our approach were also found in the colocalization analysis. This discrepancy arises from the fact that the colocalization approach only considers pairs of disease both associated at *P*<5×10^-8^ with genetic variants in overlapping credible sets. Our approach allows associations not significant at *P*<5×10^-8^ to be retained (**Extended Data Fig. 2**).

## Discussion

As genetic risk factors are becoming increasingly relevant to various fields of medicine, the ability to evaluate their impact on disease burden is crucial. In this study we take steps in this direction by presenting a new approach to quantify the effect of various genetic exposures in terms of DALYs (“lost healthy life years”) by combining genetic data with DALY estimates from the Global Burden of Disease study. The results of our main analyses can be freely explored at: https://dsge-lab.shinyapps.io/daly_genetics/.

Overall, rare deleterious variants tended to have higher effects on DALYs than common variants at the *individual* level. We also found that genetic exposures increasing the risk of ischemic heart disease tended to be most impactful in terms of DALYs, since it accounts for the largest share of population DALYs in the GBD for Finland 2019 (11.5%) (**ST3**). Also, we have shown that the *population* level impact of some common variants is comparable to important classic risk factors, such as low physical activity and diet high in sodium. Polygenic scores, especially those that capture multiple diseases (e.g., lifespan^32^ and multisite chronic pain^33^), have a sizable impact both at *individual* and *population* level (**ST13**).

The largest effects on *individual* DALYs were observed for carriers of high penetrance deleterious rare variants in *BRCA1* (breast and ovarian cancer), *BRCA2* (breast, ovarian, liver and prostate cancer), *MYBPC3* (cardiomyopathy and myocarditis), *LDLR* (ischemic heart disease) and *MLH1* (colon and rectum cancer). However, due to the rarity of these variants, the *population* impact was at most 21 yearly population DALYs per 100,000 for *BRCA2* (**ST15**), which is substantially lower than for the top common variant rs7859727 (*CDKN2B-CDKN2A*) where minor allele carriers account for 447 yearly population DALYs per 100,000.

PGSes had a moderate to high impact both at the individual and population level. The examined PGSes tended to moderately increase the risk of many diseases, with median 16 out of 80 diseases retained per PGS. Overall, the top PGSes exert their effect through cardiometabolic traits (e.g., through ischemic heart disease for shorter lifespan^32^, coronary artery disease^40^ and type 2 diabetes^41^ PGSes) or pain/addiction-related traits (e.g., through low back pain, substance use disorders, lung cancer and COPD for multisite chronic pain^33^, lower educational attainment^34^, major depressive disorder^42^, and smoking initiation^43^ PGSes). Note that the effect estimates for PGSes depend on the cutoff used. For the shorter lifespan PGS, if we instead used top 1% vs rest or top 50% vs rest cutoffs, the individual DALYs would have been 5.60 and 2.76 respectively, instead of the reported 3.81 for top 10% *vs* rest. We also note that the effect of PGSes is in part a function of the predictive performance of each PGS, so their relative importance can change and effect on DALYs will likely increase as larger GWASes are used to construct them.

Most common variants with the highest effect on DALYs affected ischemic heart disease risk (e.g., rs3798220, *LPA*; rs11591147, *PCSK9*; rs1537371, *CDKN2B-CDKN2A*), and some affected risk of dementia (rs429358, *APOE*), prostate cancer (rs183373024, *POU5F1B*), or type 2 diabetes (rs117361510, *JPH2*). The number of DALYs for a disease in the GBD is driven by how common it is, how much premature death it causes (years of life lost, YLL), and how much and for how long it lowers quality of life (years lived with disability, YLD). Common diseases that either lead to premature mortality (high YLLs e.g., ischemic heart disease) and/or long periods of living with disability (high YLDs, e.g., low back pain) account for a large number of population DALYs (**ST3**). Consequently, genetic exposures increasing the risk of high-DALY diseases predominated the results. Some variants might affect DALYs by modifying intermediate risk factors such as BMI and blood pressure and, in our analyses, we have included several variants that were associated with major risk factors. Nonetheless, we noticed that most of the highest-ranking variants in terms of DALYs are associated directly with the diseases rather than with intermediate modifiable risk factors (rs8040868 in *CHRNA5/A3/B4* being a notable exception due its impact on smoking behavior).

There are multiple strengths and limitations to our study. One key strength is that the genetic associations were estimated using individual-level data from two large biobank studies with long registry-based follow-up. We apply a shrinkage procedure to the associations, and thus obtain a conservative overview of the pleiotropic effect of genetic exposures on 80 major disease accounting for 83.1% of the total DALYs in Finland through all noncommunicable diseases in 2019^19^. In quantifying the disease burden we combine the association results with population DALYs from the GBD^19^ which reports perhaps most accurate and unbiased estimates of population-level disease burden. This is important for many outcomes for which defining the absolute disease burden (e.g., incidence) through registry data is biased downwards. For example, quantifying the number of individuals suffering from migraine using hospital episode statistics would lead to a serious underestimate (**Extended Data Fig. 8**). One important aspect of the DALY estimates from the GBD is that the disease definitions are not overlapping and the DALYs are comorbidity-corrected (see Supplementary Appendix Section 4.9 of GBD 2019^19^). Thus, double counting DALYs due to comorbidity is avoided. Finally, compared to similar studies that consider modifiable/environmental risk factors, genetic exposures are less likely to be impacted by confounding and other biases and thus more readily allow for a causal interpretation.

In **ST10** we provide an overview of the study limitations, some of which can be overcome in future iterations of this work. Here we discuss perhaps the most important ones. First, we take it as given that the DALYs estimated by the GBD^19^ are accurate and that DALYs are a meaningful measure of disease burden (which has been debated^44^). Second, the DALYs for individuals with a disease is assumed to be the same among those with and without the genetic exposures (e.g., individuals with and without a damaging *BRCA1* mutation that develop breast cancer accumulate DALYs similarly). Third, we used DALYs estimated for Finland 2019 and estimated genetic associations using data between 1972 and 2020 in Finland and United Kingdom, so the estimated effects on lifetime individual DALYs for someone born today can change as the disease incidence, medical care, and mediating factors (e.g., smoking) change. Also, we currently do not model gene-environment effects or account for age-varying effects. Fourth, although we suggest a causal interpretation to the attributable DALYs, there are important caveats to be made. Despite rigorous statistical fine-mapping, the reported variants with the highest posterior probability might tag underlying causal genetic variation. For example, the rs3798220 (*LPA*) variant is known to tag copy number variation of the Kringle IV like domain 2 in the *LPA* locus, which is the probable mechanism behind the association of rs3798220 with ischemic heart disease^45^. Another caveat lies in the possibility that a reported variant does not tag one causal variant, but multiple causal variants in LD. However, using eQTL data, previous work on quantitative lipid traits has shown that a minority of pleiotropic effects at a given locus are explained by this configuration^46^. Finally, note that we mainly present attributable DALYs for both sexes in aggregate, which might be misleading for exposures with sex-specific effects. For example, rs183373024 (*POU5F1B*) affected DALYs only through prostate cancer and benign prostatic hyperplasia (0.54 DALYs in aggregate, 1.06 DALYs in males, **ST11**).

The presented framework can be applied to other countries and ancestries under certain assumptions. First, the effect sizes of the genetic exposures need to be generalizable to the target population. There is increasing evidence that many causal variants have similar effects across continental ancestry groups^47–51^, but this does not apply to polygenic scores^52^. Assuming effect sizes are consistent, one needs to know the frequency of the genetic exposures in the target population, which is particularly challenging in countries with a heterogeneous ancestry composition. In the absence of representative genetic surveys, it might be possible to use self-reported ancestry information combined with frequency datasets such as gnomAD^53^.

In conclusion, we have proposed an approach to combine genetic association results from large biobank studies with disease burden estimates from the GBD and provided an overview of the impact of genetic exposures on lost healthy life years (DALYs). We have shown that some common variants account for as many healthy life years as some well-established modifiable risk factors. We emphasize that beyond estimating the direct impact of *genetic* exposures on DALYs, genetic results can be used to better estimate the causal effect of *modifiable* risk factors on disease burden using techniques such as Mendelian randomization^54^. While genetic risk factors are not yet modifiable in practice, novel approaches based on *in vivo* gene editing are being developed^16^ and some have been tested in clinical trials^17, 18^. Estimating the total impact of genetic risk factors on disease burden might help prioritize these types of interventions.

## Methods

### General statistical methods

All analyses were performed using R version 4.0.3. All *P*-values are nominal. Error bars represent 95% confidence intervals. Confidence intervals for the disease-specific DALYs were generated using the delta method. Confidence intervals for total DALYs for each exposure were estimated by repeating 10,000 times the computation of total DALYs while resampling the log-HR estimates from a multivariate normal distribution corresponding to the approximate sampling distribution of log-HRs, and by taking the 2.5^th^ and 97.5^th^ percentiles as the bounds for the 95% confidence intervals (**Supplementary Information**). The sex-specific results were generated through estimating the HRs stratified by sex and using sex-specific population DALYs reported by the GBD^19^. For all error estimates we assumed there was no uncertainty in frequencies of the exposures (e.g., allele frequencies) or DALYs reported by the GBD, so we only consider variation from HR estimation in the error estimates.

### Participants

FinnGen and UK Biobank^55^ are large-scale population-based prospective cohort studies from Finland and the United Kingdom, both with phenotypic data ascertained from registries (**Fig. 1**). In FinnGen, we used participants from the Finnish ancestry in data freeze 7 with n=309,136 (56.2% female) with median (IQR) age at start of follow-up of 15.1 (0-26.3) and at end of follow-up 62.2 (47.1-72.9). The median (IQR) follow-up length was 47.3 (41.6-47.7) years (**ST1**). Follow-up for FinnGen started on January 1^st^, 1972 and ended on August 31^st^, 2020. For UKB we used participants of European ancestry n=426,612 (54.1% female) with median (IQR) age at start of follow-up of 47.2 (39.5-52.4) and at end of follow-up 69.1 (61.7-74.2). The median (IQR) follow-up length was 22.3 (22.3-22.3) years (**ST1**). Follow-up for UKB started on January 1^st^, 1998 and ended on April 30th, 2020.

### Disability-adjusted life years

As a measure of disease burden we used the disability-adjusted life years (DALYs) from the 2019 Global Burden of Disease study (GBD)^19^ with data publicly available at http://ghdx.healthdata.org/. DALYs are a metric for measuring population-level disease burden that combines a measure of premature mortality called *years of life lost* (YLL) and a measure of healthy life years lost due to lowered quality of life called *years lived with disability* (YLD), so DALYs are the sum of YLDs and YLLs (**Extended Data Fig. 1**). The GBD is a longitudinal study which estimates incidence, prevalence, mortality, YLLs, YLDs and DALYs due to various collectively exhaustive and mutually exclusive diseases and injuries (369 in 2019) for hundreds of countries (204 in 2019)^19^. GBD strives to model unbiased estimates using various sources of information, including census data, household surveys, civil registration and vital statistics, disease registries, health service use data, and more. The estimation process for DALYs is complex and varies from disease to disease, see the 2019 GBD publication^19^ and its Supplementary Appendix 1 for a detailed description of the methods. Because all of our genetic exposures, except rare variants, were measured in FinnGen, we used the 2019 GBD metrics for Finland and present all estimates for Finland, although the estimates could be calculated for any country in the GBD given some assumptions. See **ST3** for a list of DALYs, YLLs and YLDs for the included diseases.

### Disease phenotypes

To define the disease phenotypes, we manually mapped 89 mutually exclusive noncommunicable diseases from the 2019 GBD to FinnGen clinical endpoints (**ST2**), of which we retained 80 based on requiring at least 500 individuals to have the disease in FinnGen. These 80 diseases account for 83.1% of population DALYs from noncommunicable diseases in Finland 2019 (**ST3**). We did not map diseases from the GBD that were too rare, not well enough captured by the registries, or include too heterogenous a set of diseases (e.g., “other cardiovascular and circulatory diseases”). Phenotyping in FinnGen relied on information on diagnoses starting from 1972 in the hospital discharge registry^56^, the causes of death registry, and the cancer registry. The drug purchases registry was additionally used starting from 1995 for selected diseases (e.g., migraine). We completely excluded diseases with less than 500 cases in FinnGen. Phenotyping in UKB was performed via groups of ICD-10 codes mapped from the FinnGen endpoints (**ST2**), relying on ICD-10 diagnosis codes from the hospital episode statistics, cancer registry, and causes of death registry data starting from 1998. The cumulative incidence of the diseases varied from 14.93% (cataract) to 0.16% (acute glomerulonephritis) in FinnGen. We analyzed the following diseases only in FinnGen as in UKB there were less than 500 cases: Hemoglobinopathies and hemolytic anemias (n=480 in UKB), Hodgkin Lymphoma (n=464), autism spectrum disorders (n=164), eating disorders (n=163), acne vulgaris (n=106), acute glomerulonephritis (n=58), attention-deficit hyperactivity disorder (n=26), and conduct disorder (n=13). As both FinnGen and UKB rely mainly on diagnosis codes recorded at hospitals, conditions that are usually managed in the primary or outpatient care setting are under-ascertained, for examples see **Extended Data Fig. 8**.

### Common variants

We provide information on genotyping, imputation, quality control, fine-mapping, and LD clumping in **Supplementary Information**. For each variant, we defined the minor allele to be the allele less common in FinnGen. Due to symmetry in attributable DALY estimation, going from 1 to 0 copies vs 0 to 1 copies of an allele only changes the sign of the individual attributable DALYs estimates. After fine-mapping, LD clumping, and shrinkage, we included a total 1044 common variants (FinnGen MAF>0.01) for the analysis (**Fig. 1**). FinnGen SuSiE fine-mapping^20^ results for all FinnGen endpoints used to define the 80 diseases (**ST2**) were used to select, for each credible set, the genome-wide significant variant (P<5×10^-8^) with the highest posterior inclusion probability, resulting in 564 variants. Additionally, we included all FinnGen coding variants with a P<5×10^-8^ association with any FinnGen endpoint (n=325). We used UKB fine-mapping results^21^ for continuous risk factor traits (BMI, systolic blood pressure, cigarettes per day, HLD-C, LDL-C, and HbA1c) and selected, for each credible set, variants with P<5×10^-12^ and the highest posterior inclusion probability, which resulted in inclusion of 155 additional variants. We labeled common variants as Finnish-enriched if they had FinnGen MAF enrichment at least 5-fold compared to the non-Finnish-non-Swedish-non-Estonian European (NFSEE) ancestry MAF and the NFSEE MAF was over 0.01. In **ST5** we report all common variants included in the analysis. We separately imputed^22^ 74 alleles in 7 HLA genes (**ST9, Supplementary Information**) for the HLA region (chr6:29691116 to chr6:3054976 in GRCh38) and determined 6 haplotypes (**ST8**) for *APOE*.

### Rare deleterious variants

To analyze the effects of rare deleterious variants on DALYs, we used whole exome-sequencing data from a subset of participants in UKB (n=174,379) from the December 2020 release. We processed the data using *Hail* version 0.2.59-63cf625e29e5. We directly processed the quality controlled *PLINK*^57^ files provided by UKB. We annotated the data using the Ensembl Variant Effect Predictor^30^ using the same approach used in gnomAD and specifically the function *gnomadvep.vep_or_lookup_vep.* VEP annotations were processed using the function *gnomadvep.process_consequences*, consistently with the gnomAD definition of pLOF, missense and synonymous variants, using the canonical transcript. We also extracted ClinVar^28^ annotated variants (accessed in November 2020) and germline variants in *BRCA1* and *BRCA2* (accessed in November 2020) annotated by the ENIGMA consortium^29^. From ClinVar we considered “pathogenic” and “likely-pathogenic” variants (no filtering on star status or number of submitters) and from ENIGMA we considered “pathogenic” variants. Variants with frequency greater than 0.001 were excluded because they are less likely to be deleterious. We considered all genes part of the the American College of Medical Genetics and Genomics (ACMG) recommendations for reporting of incidental findings in clinical exome and genome sequencing studies^27^. We formed two types of rare variant burdens for individuals for each gene: 1) the pLOF burden was set as positive if there was at least 1 variant annotated as pLOF and 2) the ClinVar/ENIGMA burden was set as positive if there was at least 1 variant annotated as “pathogenic” in ENIGMA^29^ for *BRCA1* and *BRCA2* and for other genes “likely pathogenic” or “pathogenic” in ClinVar. Due to statistical power considerations, we restricted our analysis so that at least 35 individuals had a positive burden, resulting in 9 genes for both burden types (**ST4**).

### Polygenic scores

In FinnGen, we included 30 genome-wide polygenic scores (PGS) for traits of interest constructed from publicly available summary statistics, see **ST6** for references. We selected PGSes for psychobehavioral traits (e.g. cognitive ability, neuroticism), major chronic diseases (e.g. coronary artery disease, depression), and major risk factors (e.g. LDL-C, blood pressure) to cover traits of interest with high quality summary statistics available. PGSes were only analyzed in FinnGen, since many of the original summary statistics included UK Biobank. We used PRS-CS^58^ for generating the PGSes using external LD reference panel (1000 Genomes Europeans). We used the PRS-CS-auto algorithm, which learns the model’s global scaling parameter from the data and performs well with large datasets. Default PRS-CS parameters were used and only HapMap 3 SNPs were considered (see https://github.com/FINNGEN/CS-PRS-pipeline for code). Scores for lifespan, educational attainment, cognitive performance, and intelligence were reversed before analysis to make all scores on net deleterious in terms of DALYs. For defining the exposure for survival models, we coded individuals for each PGS as 1 if they were in the top 10% of the score distribution and 0 if not. Consequently, the individual DALYs can be interpreted as the effect on lifetime DALYs if the average individual in top 10% of the PGS were to have their PGS be that of the average in the bottom 90% of the PS.

### Survival models

To estimate the HRs between all genetic exposure-disease pairs we used Cox proportional hazards regression via the *coxph* function in the *survival* package version 3.2-11. The model was additive for allele counts. Sex and first 10 principal components of population structure were included as covariates. We used calendar age as the time-scale and age at first record of the disease in the registries as time-to-event. Individuals were censored at death, emigration, or end of registry-based follow-up (August 31^st^ 2020 in FinnGen, April 30^th^ 2020 in UKB). For the common variants, the HRs were estimated both in FinnGen and UKBB separately and combined using fixed effects inverse-variance weighted meta-analysis. A comparison of effect sizes between FinnGen and UK biobank is provided in **Extended Data Fig. 3**. We did not account for left censoring, nor did we account for relatedness of the subjects due to computational limitations. For the rare variant burden analysis in UKB, we used Cox regression with Firth’s Penalized Likelihood^59^ via *coxphf* package version 1.13.1 and included sex and the 4 first genetic principal components as covariates.

For the sensitivity analysis in FinnGen accounting for relatedness in estimating standard errors for the log-HRs, we generated a family indicator from genotype data using KING^60^ version 2.2 only including HapMap 3 variants. Individuals up to 3^rd^ degree of relatedness were included in the same family. The robust standard errors were estimated using a family indicator to compute robust standard errors by including *cluster(family_id)* as a covariate in *coxph*.

### Shrinkage

We use prior information to apply a shrinkage procedure to the HRs for exposure-disease pairs to reduce the effect of sampling variation on our results. Denote by *b_e,d_*, the log-HR of genetic exposure *e* on disease *d=1,..,80.* One genetic exposure at a time, we set the prior distribution of *b_e,d_*, to be a mixture distribution between the point mass at 0 and a 50:50 mixture of two normal distributions with means at −0.3 and 0.3, respectively, and with a standard deviation of 0.1. We denote the mixture weight of the non-zero component by *p_e_*, which can be interpreted as the exposure-specific proportion of non-zero effects across the 80 diseases. We set the prior distribution of *p_e_* to a Beta(α = 1, β = 19) distribution, that has an expected value of 0.05. The full probability model is

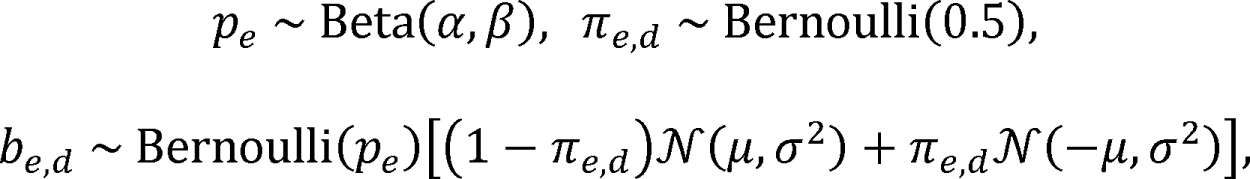

where α = 1, β = 19, μ = 0.3 and σ = 0.1.

This model implies that, before seeing the data, for each genetic exposure, we expect a non-zero effect for 4 (= 0.05 x 80) diseases and the direction of the non-zero effects are equally likely to be risk increasing (centered around HR = 1.35) as protective (centered around HR = 0.74). In practice, which effects are shrunk to zero and which are retained as non-zero, does not vary considerably when these prior parameters are varied (**Extended Data Fig. 6**). For each genetic exposure *e* at a time, we use a Markov Chain Monte Carlo procedure with 10,000 iterations (**Supplementary Code**) to estimate the posterior probabilities of the log-HRs (*b_e,d_*) for diseases *d*=1,2,…,80 coming from the null model (point mass at zero). We discard any log-HRs where the null probability is over 10%, and, for the remaining log-HRs, we use the maximum partial likelihood estimates from the Cox proportional hazards model in the downstream analyses.

### Attributable disability-adjusted life years

Similarly to the GBD^9^, we use the HRs and frequencies of the exposures to estimate attributable DALYs one disease at a time. We use the multilevel exposure formula^61^ for the population attributable fraction (i.e., the fraction of cases of disease *d* caused by the exposure levels in the population deviating from counterfactual levels):

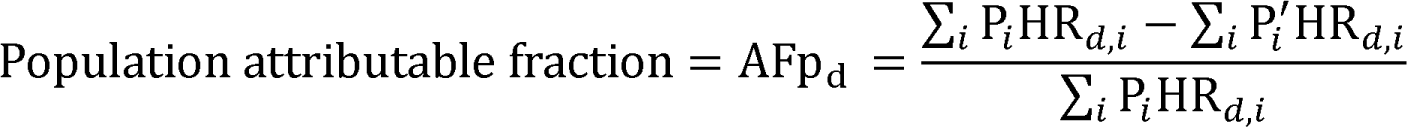

where HR*_d,i_* is the hazard ratio for disease *d* at exposure level*i* (e.g., 1 copy) relative to reference (e.g., 0 copies) and P*_i_* is the fraction of the population at exposure level *i* and P’*_i_* represents the fraction of the population at exposure level *i* in the counterfactual scenario (e.g., if all with 1 copy instead had 0: P’*_1_* =0, P’*_0_* = P_0_ + P_1_, and P’*_2_* = P_2_).

As an example, for individuals carrying 0, 1, and 2 alleles with respective population frequencies of P_0_ = 0.7 P_1_ = 0.2 and P_2_ = 0.1 and HRs for disease *d* of HR_d,0_ = 1.00 HR_d,1_ = 1.35, and HR_d,2_ = 1.82. For calculating attributable DALYs from individuals carrying 1 vs 0 copies of the allele, define the counterfactual frequencies as P^’^_0_ = 0.9, P^’^_1_ = 0.0, and P^’^_2_ = 0.1 (i.e., making all with 1 copy have 0 copies instead). Plugging these numbers into the AFp formula produces the population attributable fraction of disease cases from those carrying 1 vs 0 copies of the allele (the fraction of cases that would be prevented if all with 1 allele had instead 0), which is 0.061 in this case, so approximately 6.1% of disease cases would be prevented if all with 1 copy had instead 0 copies at birth.

We then assume that the estimated population attributable fraction of disease cases can be interpreted as the population attributable fraction of DALYs, which is true if all disease cases contribute on average the same amount of DALYs independent of whether they have the genetic exposure or not. This assumption does not hold if, for example, deleterious *BRCA1* mutation carriers develop breast cancer earlier and consequently accumulate more DALYs per case. Then, multiplying the population attributable fraction of DALYs (AFp_d_) by the population DALYs per year per 100,000 reported by the GBD (DALY_d_) gives the *population attributable* DALYs, interpreted in our example as *the expected loss of healthy life years per year per 100,000 if the population frequencies of the exposure were* P^’^_i_ *instead of* P_i_ (in our example, all with 1 copy had instead 0 copies).

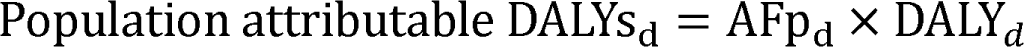

We further estimate *individual attributable DALYs* for binary counterfactuals (e.g., having 1 vs 0 copies, being in top 10% of a PS vs bottom 90%) by dividing the *population attributable* DALYs per 100,000 by the number of individuals with the exposure out of 100,000 (100,000 x P_i_) and multiplying by life-expectancy at birth (L):

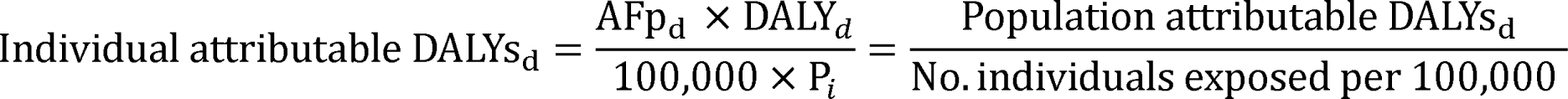

where DALY_d_ represents the population DALYs per year per 100,000 through disease *d* from GBD, P*_i_* is the fraction of population for which the exposure is changed (e.g., fraction of those with 1 copy), and L represents the life-expectancy at birth (which is included to convert yearly DALYs into lifetime estimates). These individual DALYs are interpreted as *the expected loss of healthy life years for an individual caused by having the genetic exposure at birth*. Finally, both population attributable DALYs and individual DALYs can be summed up across the 80 diseases to arrive at the total impact of the genetic exposure. See Witte et al.^62^ for discussion on how population attributable fractions relate to other measures of genetic contribution.

## Data availability

We present all attributable DALY results in **ST8, ST9**, and **ST11-ST16**. Results for common variants, HLA alleles, PGSes can be explored through plots at https://dsge-lab.shinyapps.io/daly_genetics/ Code for central parts of analyses can be found at https://github.com/dsgelab/dalys_code

## Data Availability

All summary statistics level data are available in the supplemental tables.

## Acknowledgements

We thank the whole FinnGen team for their contribution. In particular, we thank Mari K. Niemi for methodological support and comments to the manuscript, Samuli Ripatti and Masahiro Kanai for insightful comments on the manuscript, Pietro della Briotta Parolo for constructing the polygenic scores and kinship information in FinnGen. We also thank John J. McGrath (Aarhus University) for very helpful comments on the manuscript. S.J. was supported by the Academy of Finland (grant no. 341747). A.G. was supported by Academy of Finland (grant no. 323116) and by the European Research Council (ERC) under the European Union’s Horizon 2020 research and innovation programme (grant no. 945733). This project has also received funding from the European Union’s Horizon 2020 research and innovation programme under grant agreement no. 101016775. The FinnGen project is funded by two grants from Business Finland (HUS 4685/31/2016 and UH 4386/31/2016) and nine industry partners (AbbVie, AstraZeneca, Biogen, Celgene, Genentech, GSK, MSD, Pfizer, and Sanofi). Following biobanks are acknowledged for collecting the FinnGen project samples: Auria Biobank, THL Biobank, Helsinki Biobank, Northern Finland Biobank Borealis, Finnish Clinical Biobank Tampere, Biobank of Eastern Finland, Central Finland Biobank, Finnish Red Cross Blood Service Biobank. UKB analyses were conducted under application no. 31063.

## Author contributions

A.G. conceptualized the study. A.G. and S.J. designed the analysis plan. S.J. performed most of the analyses that were not part of the FinnGen GWAS pipeline. The manuscript was written by S.J. and A.G. The phenotyping approach for FinnGen was mainly developed by T.K. and A.H., and they also provided methodological support. J.K. participated in genetic fine-mapping and provided methodological support. M.C., J.T.R., N.M., and K.E.S. provided methodological support. H.M.O. provided advice on the HLA analyses. M.P. provided statistical support by helping develop the approach for quantifying uncertainty via resampling and helping develop the shrinkage approach. All authors participated in reviewing the manuscript.

## Extended Data Figures

**Extended Data Fig. 1:**
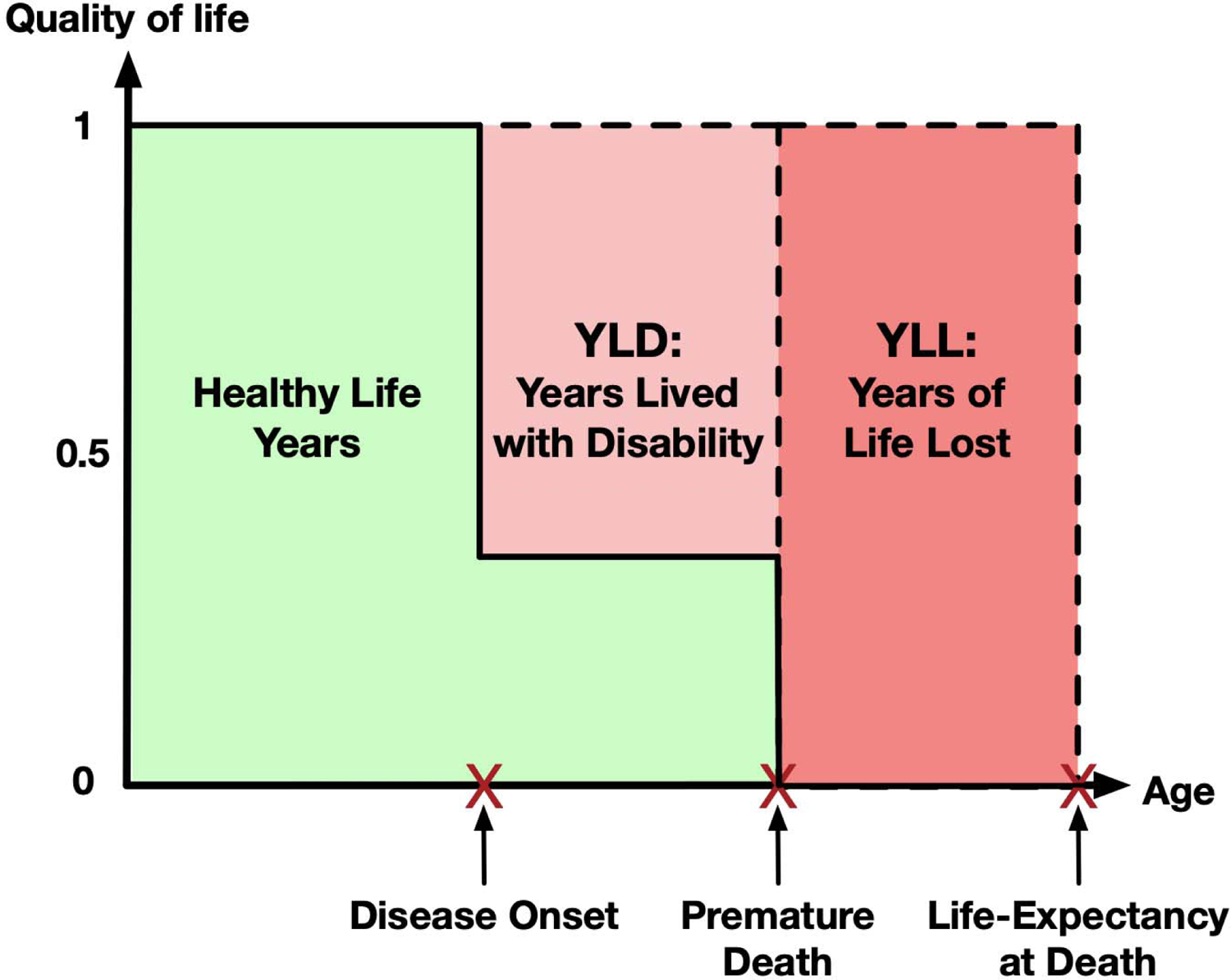
Schematic representation of how disability-adjusted life years (DALYs) are constructed from years lived with disability (YLD), and years of life lost (YLL). DALYs are a metric for measuring population-level disease burden that combines a measure of premature mortality called years of life lost (YLL) and a measure of healthy life years lost due to lowered quality of life called years lived with disability (YLD), so DALYs are the sum of YLDs and YLLs.

**Extended Data Fig. 2:**
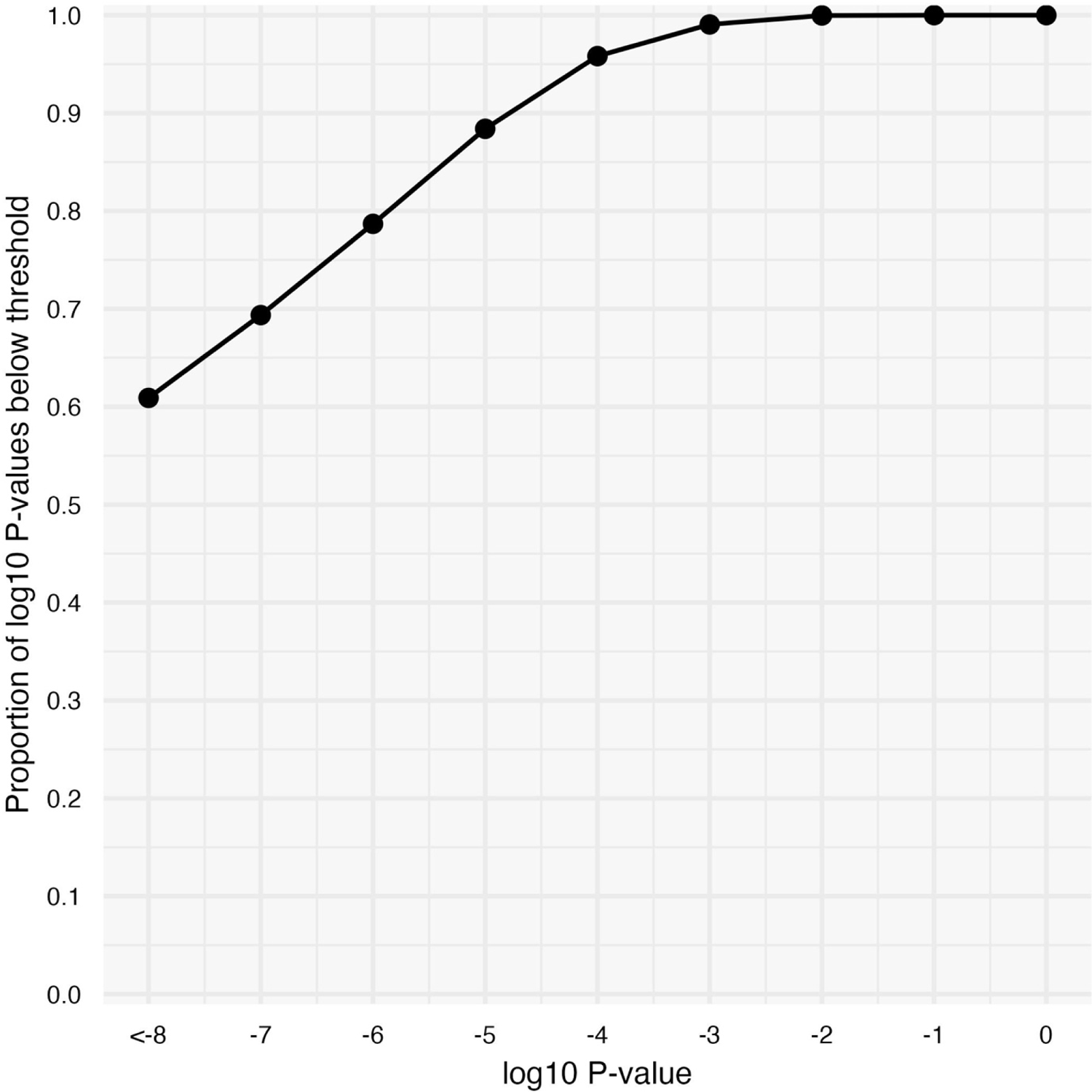
Cumulative distribution plot of log_10_ P-values for all reported HRs between genetic exposures and diseases (n=3,123). 67.1% of the associations were genome-wide significant (P<5×10-8) and 99% had an association with P<7.3×10-4

**Extended Data Fig. 3:**
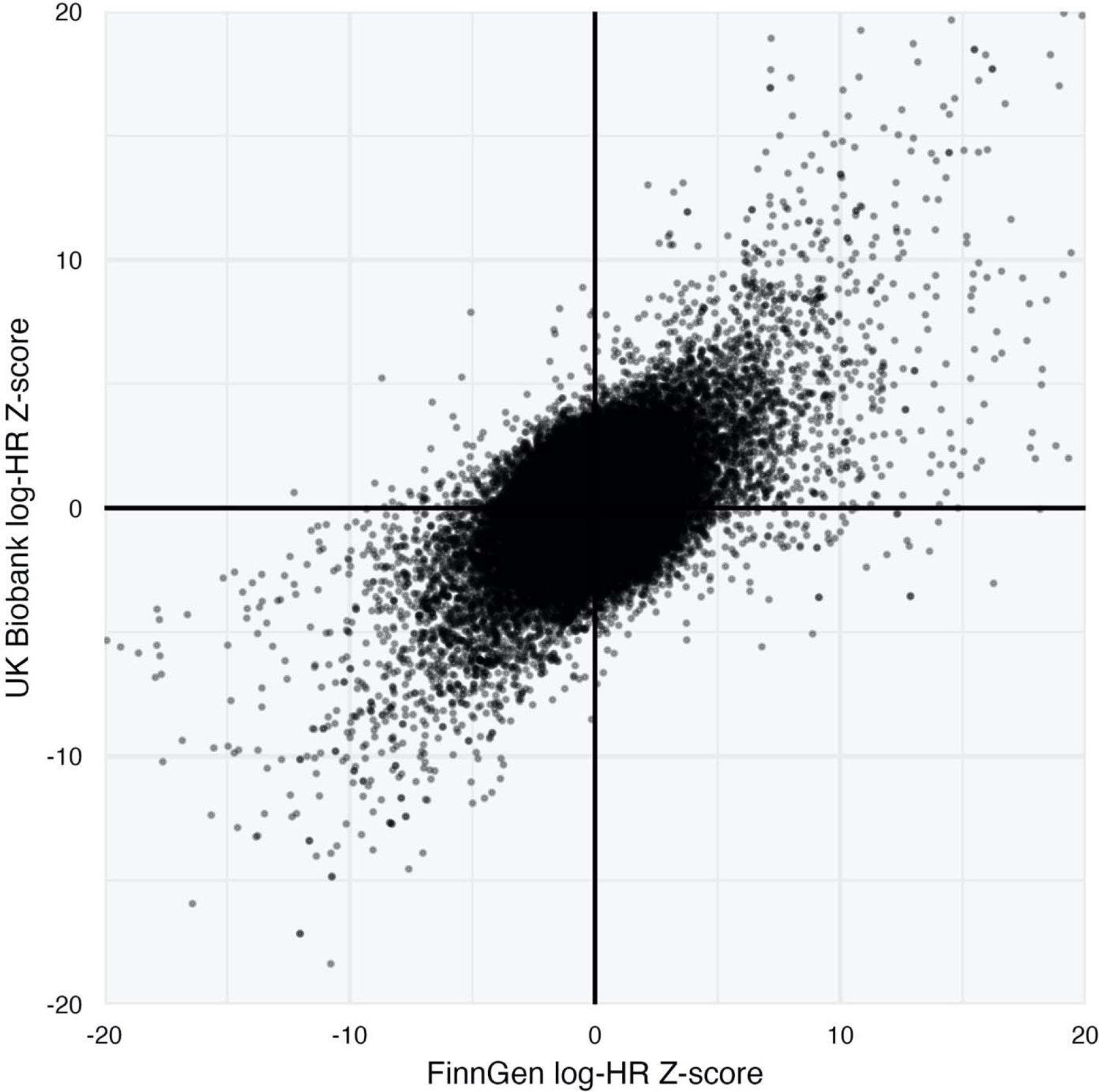
Comparison of effect sizes (log-HR Z-score) for common variant-disease associations (n=68,616) estimated both in FinnGen and UK Biobank. Axes are truncated at ±20, leaving out 36 points.

**Extended Data Fig. 4:**
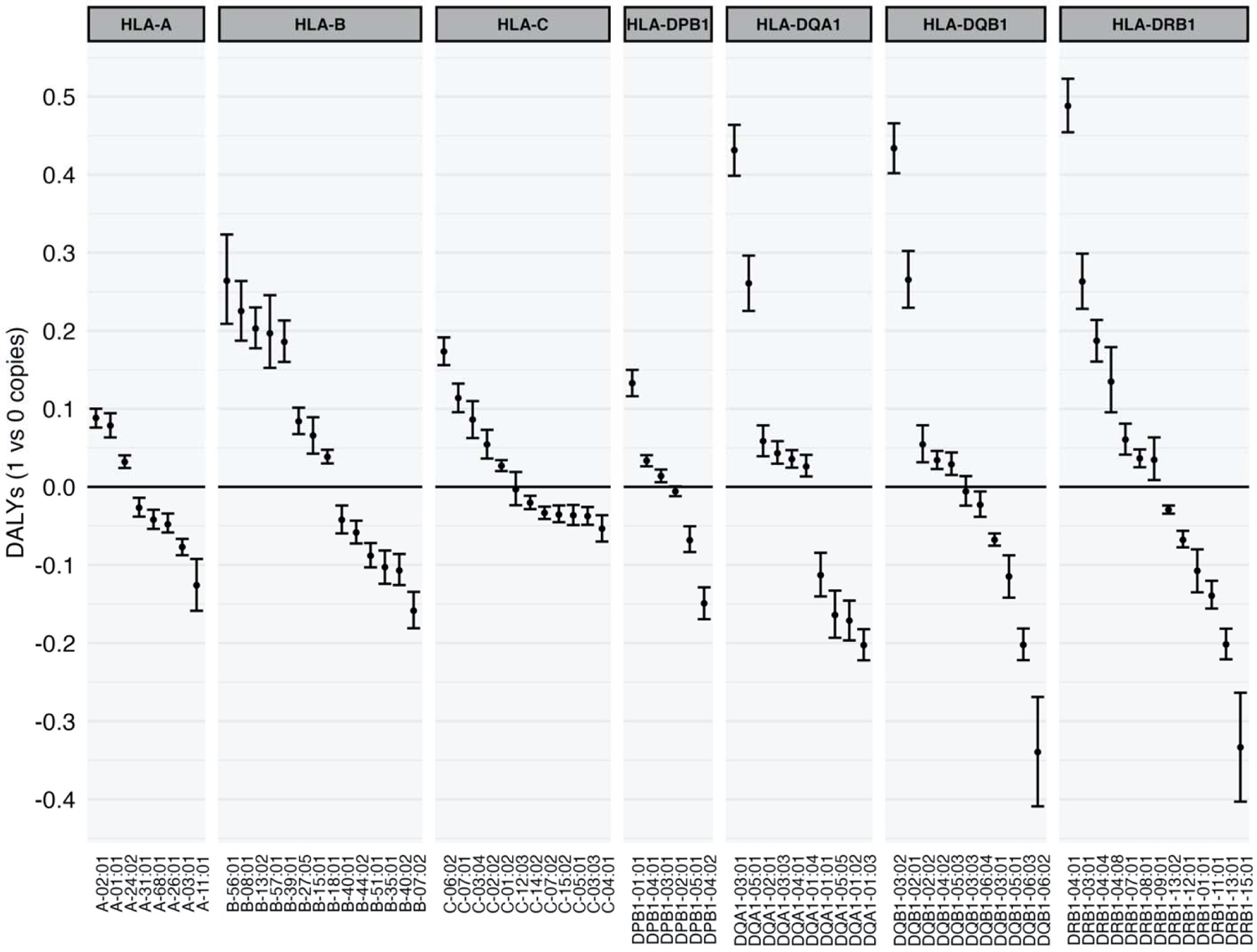
Effect of HLA alleles on DALYs by HLA gene. Note that each gene is multiallelic, so the comparison of “1 vs 0 copies” is more precisely the effect for having “1 copy of the allele in question and 1 average copy of other allele” vs “having 0 copies of the allele in question and 2 average copies other alleles”. See ST9 and Supplementary Information for details. Error bars denote 95% confidence intervals.

**Extended Data Fig. 5:**
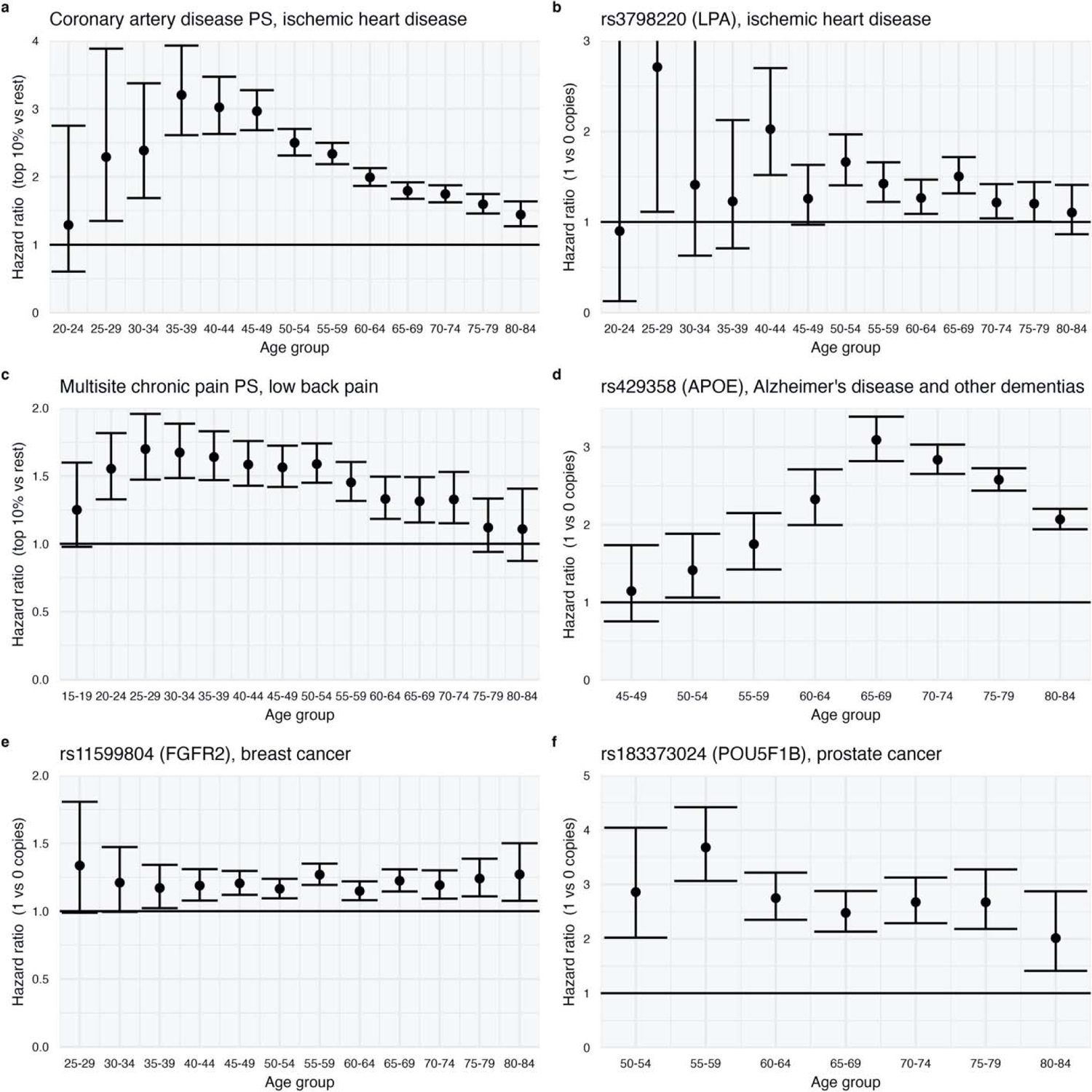
Hazard ratios by age group in FinnGen. For each age group we estimated the HRs via a Cox proportional hazards model including individuals whose follow-up started prior to beginning of the age interval that did not have a previous record of the disease. The figure demonstrates age-varying HRs especially for ischemic heart disease, Alzheimer’s disease and other dementias, and low back pain (a,b,c,d). For breast cancer and prostate cancer, the HRs were approximately constant across age groups (e,f).

**Extended Data Fig. 6:**
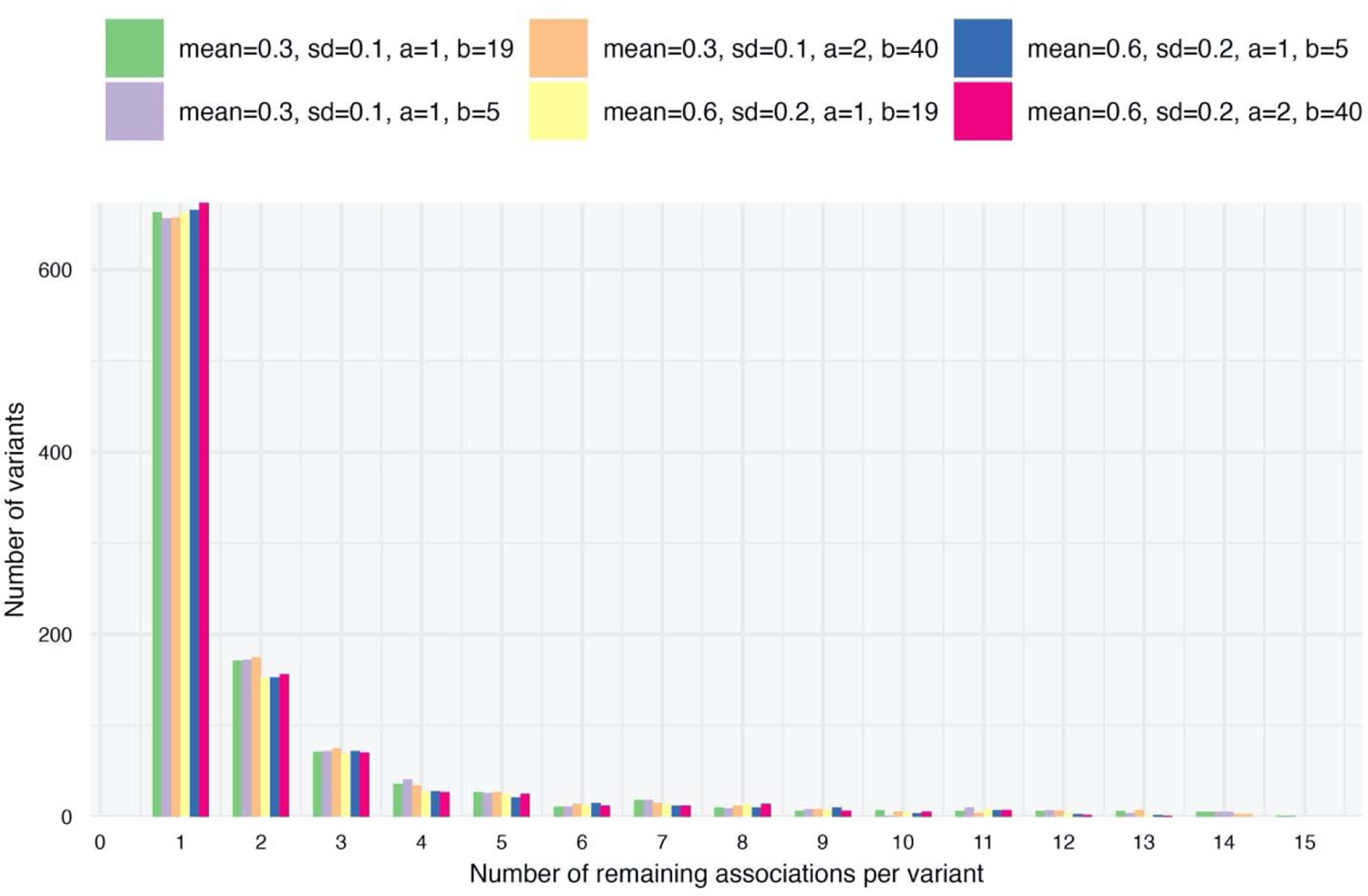
Histogram of the number of retained disease associations after shrinkage using different prior parameters. In this plot we only consider common variants. “mean” and “sd” correspond to the μ and σ prior parameters, “a” and “b” correspond to the α and β prior parameters (Methods). Overall, the distribution of retained association is not sensitive to the choice of prior parameters.

**Extended data Fig. 7:**
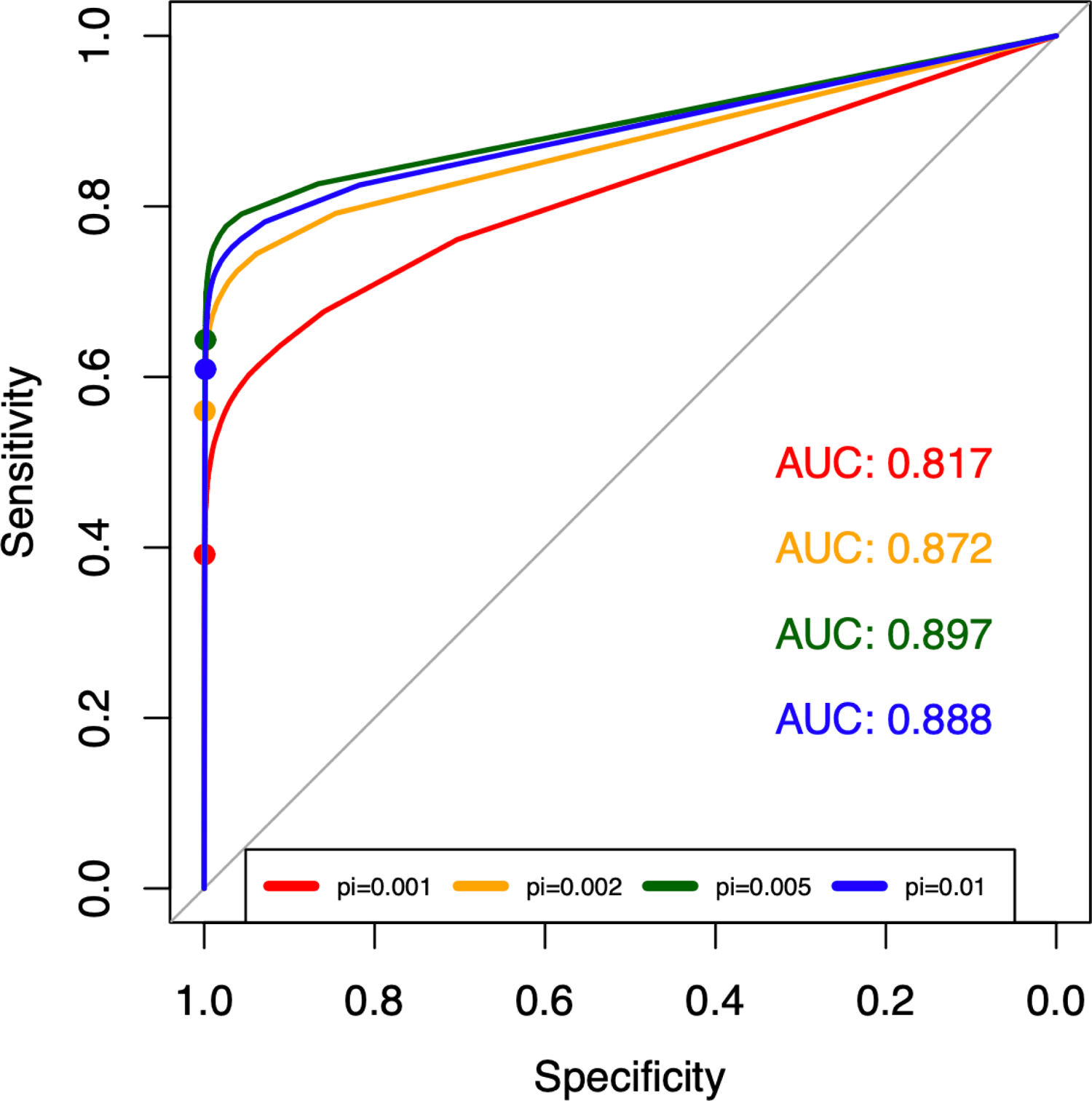
Receiver operating characteristic curves using our shrinkage method a a classifier to identify true causal variants in simulated data. Using Hail, we simulated GWA summary statistics for 80 binary phenotypes with heritability sampled uniformly between 0. and 0.6. We repeated this for four scenarios where the probability of a variant being causal (pi is 0.001, 0.002, 0.005, and 0.01 (Supplementary Information). We then applied our shrinkag procedure and classified those variants with posterior probability of being from the null model of <10% as being causal (colored dots on lines represent this threshold). See ST7 for details o classification performance.

**Extended data Fig. 8:**
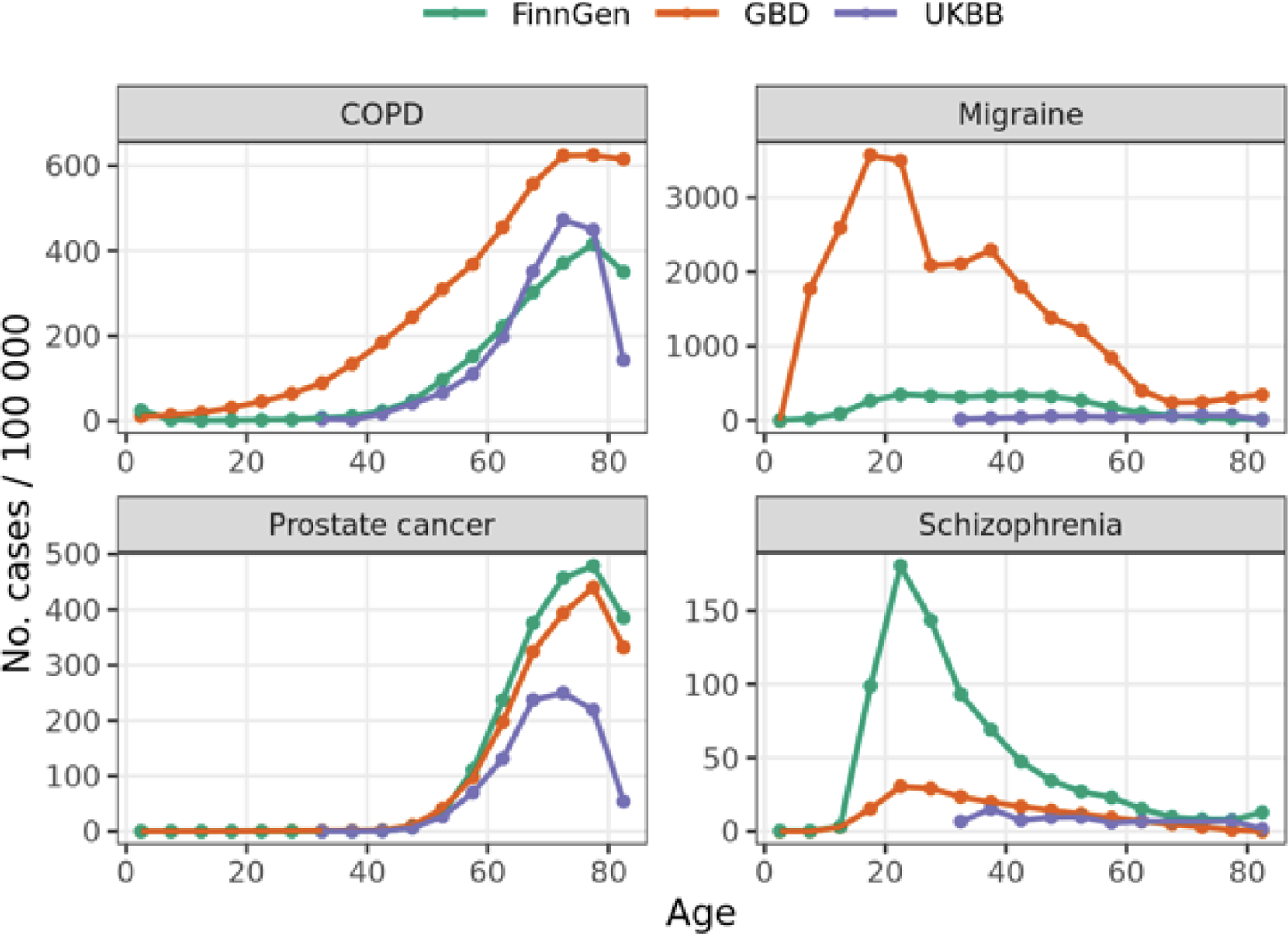
5-year incidence estimates of COPD, migraine, prostate cancer, and schizophrenia in FinnGen and UK Biobank (UKBB) compared to the population incidence estimates from the Global Burden of Disease Study 2019 for Finland 2019 (GBD). Number of new cases per 100,000 was calculated as number of new cases per 100,000 during each age interval divided by person-years contributed by individuals that were not cases in the earlier age intervals. The incidence of some diseases like prostate cancer is well captured in UKB and FinnGen, while other disease, such as migraine, as severely under-estimated.

