## Supplementary Information for "Genetic risk factors have a substantial impact on healthy life years"

### Cohort and data sources description

#### FinnGen

FinnGen is a public-private partnership research project combining genotype data generated from Finnish biobanks and digital health record data from Finnish health registries (<https://www.finngen.fi/en>) aiming to provide new insight in disease genetics. Participants for FinnGen include participants of legacy cohorts, both population-based epidemiological cohorts (initiated as far back as 1992) and disease-based cohorts, and volunteers from biobanks. All samples donated to biobanks are eligible for FinnGen. Information about the opportunity to donate a sample and medical information for biobank research was distributed by leaflets, advertisement campaigns in press and on TV, and by dedicated biobank nurses in hospitals. In this analysis we used participants passing genotyping quality control from the Finnish ancestry from data freeze 7 with n=309,136. Descriptive statistics are provided in **ST1**.

Like other Nordic countries, Finland has nationwide electronic health registers originally established primarily for administrative purposes to monitor healthcare usage^1^. These registers cover virtually all major health-related events, such as hospitalizations, prescription drug purchases, medical operations, cancers, and deaths. All registers can be linked using the unique personal identity code which is given to every permanent resident of Finland (<https://dvv.fi/en/personal-identity-code>). In this context, loss to follow-up can only result from emigration.

Registry data on all participants was collected from different national registers, including hospital and outpatient visits in HILMO - Care Register for Health Care, AvoHILMO - Register of Primary Health Care visits (diagnoses: ICD 8,9,10; operations: NOMESCO Classification of Surgical Procedures), Causes of Death (immediate, contributing, and underlying causes of death on the death certificate with ICD codes), reimbursed medication entitlements and prescription medication purchases (using ATC codes and specific Social Insurance Institution of Finland reimbursement codes), and the Finnish Cancer Registry (using ICD-O-3 codes). The diagnostic accuracy and validity of these registers has been reviewed in multiple previous publications, for examples see^2–4^, for reviews see^5,6^.

#### UK Biobank

We additionally included participants from the UK Biobank study (UKB), which is a population-based cohort study that recruited around 500,000 people aged between 40 and 69 years from 2006 to 2010 across the United Kingdom. For details on participants and recruitment, see Sudlow 2015^7^ and Bycroft 2018^8^. Descriptive statistics are provided in **ST1**. In this analysis we used participants of European ancestry passing genotyping quality control (n=426,612), using recorded ICD-10 diagnoses from death registrations, cancer registrations, and hospital inpatient episodes to define disease phenotypes.

### Disease outcomes definitions

#### FinnGen

To define similar disease phenotypes in FinnGen as those in the Global Burden of Disease study, we manually mapped 89 important medical conditions from the Global Burden of Disease study 2019^9^ (GBD) into FinnGen clinical endpoints definitions (see <https://www.finngen.fi/en/researchers/clinical-endpoints> for the definitions or search in <https://risteys.finngen.fi/> to explore different endpoints). The mapping (**ST2)** was performed by matching the ICD-10 definitions in the GBD as closely as possible to existing FinnGen clinical endpoints (or combinations of FinnGen endpoints, separated with “|” operator in **ST2**).

We did not attempt to map infectious diseases, accidents, injuries, sufficiently rare conditions, conditions not ascertainable through registries, or conditions for which the GBD 2019 does not report DALYs for Finland. Out of these 89, we further removed conditions for which there are under 500 cases among the subjects, ending up at 80 medical conditions which account for 83.1% of total population level yearly DALYs from noncommunicable diseases in Finland 2019 (**ST3**).

For example, ischemic heart disease definition in GBD through causes of death encompasses ICD-10 codes I20-I25.9, but our definition of ischemic heart disease through FinnGen consists of combining 5 FinnGen endpoints: I9_MI (myocardial infarction, IC10-codes I21-I22, ICD-9 and ICD-8 code 410), I9_MI_COMPLICATIONS (complications following myocardial infarction, ICD-10 code I23), I9_POSTAMI (status post acute myocardial infarction, ICD-10 code I25.3, ICD-9 and ICD-8 code 412), I9_CORATHER (coronary atherosclerosis, ICD-10 codes I24-I25, T82.2, Z95.1), and I9_REVASC (coronary revascularization, defined through NOMESCO operation codes for coronary angioplasty and coronary artery bypass grafting). The first occurrence of any of these five endpoints was coded as the event of ischemic heart disease.

We did not define disease phenotypes just using the ICD-10 codes provided in GBD for multiple reasons. First, ICD-10 based definitions for all disease conditions in GBD are not available (e.g., low back pain). Second, even when GBD presents ICD-10 diagnoses, as follow-up for FinnGen for this study started in 1968, diseases need to also be defined through ICD-8 (in use until Dec 1987) and ICD-9 (in use from Jan 1987 to Dec 1995). Third, the FinnGen clinical endpoint phenotyping algorithms have been hand-crafted to utilize various sources of registry data in addition to diagnosis codes (e.g., operation codes, prescription drug purchases, special drug reimbursement codes for certain diseases).

For example, migraine is a disease commonly managed in the primary care setting, so relying on diagnosis codes present in hospital and secondary care specialist visits data will lead to serious under-ascertainment of migraine. Also, there is misclassification of other types of headaches as migraine. The FinnGen endpoint we used to define migraine (MIGRAINE_TRIPTAN) requires that a patient has at least one purchase of prescribed triptans, capturing for example patients that do not have a diagnosis code for migraine in the available registries that do not include primary care.

#### UK Biobank

In order to make disease definitions in UK Biobank comparable to those in FinnGen, for each FinnGen definition of a disease, we considered the ICD-10 codes used in the FinnGen definition and through use of regular expressions assign UK Biobank participants diagnosed with appropriate ICD-10 codes in either the hospital episode statistics data, the cancer registry, or the causes of death registry as having the disease. The ICD-10 codes and regular expressions used to define disease outcomes in UK Biobank are provided in **ST 2**. For UK Biobank, out of the 80 diseases examined in FinnGen, we ignored diseases with less than 500 cases to avoid issues in fitting the Cox models, resulting in 72 diseases.

### Genotyping, Imputation and quality control

#### FinnGen

Samples for FinnGen were genotyped using Illumina and Affymetrix arrays (Illumina Inc. San Diego, and Thermo Fisher Scientific, Santa Clara, CA, USA). Genotype calls were made with GenCall or zCall for illumina and the AxiomGT1 algorithm for Affymetrix data. Chip genotyping data produced with previous chip platforms and reference genome builds were lifted over to build version 38 (GRCh38/hg38) following the protocol described in [dx.doi.org/10.17504/protocols.io.xbhfij6](https://dx.doi.org/10.17504/protocols.io.xbhfij6). Participants with ambiguous sex, genotype missingness of over 5%, excess heterozygosity (±4 SD) and non-Finnish ancestry were excluded. Variants with over 2% missingness, low Hardy-Weinberg equilibrium *P*-value (<10^-6^) and minor allele count under 3 were excluded. Array data pre-phasing was performed with Eagle 2.3.527 (<https://data.broadinstitute.org/alkesgroup/Eagle/>) with default parameters, except the number of conditioning haplotypes was set at 20,000. Genotype imputation was performed with Beagle 4.128 as described in [dx.doi.org/10.17504/protocols.io.xbgfijw](https://dx.doi.org/10.17504/protocols.io.xbgfijw) by using the SISu v3 reference panel developed from data on 3775 high-coverage (25-30x) whole-genome sequenced Finns.

#### UK Biobank

For detailed information on genotyping, imputation and quality control for the UK Biobank data see Bycroft 2018^8^. The genotyping was performed using the Applied Biosystems UK BiLEVE Axiom Array or the Applied Biosystems UKB Axiom Array. The genotype imputation was performed using a combination of the Haplotype Reference Consortium, UK10K and 1000 Genomes Phase 3 reference panels by IMPUTE4 software. We excluded variants with INFO score ≤ 0.8, MAF ≤ 0.01 and Hardy–Weinberg equilibrium P value ≤ 1×10^−10^.

### Principal components and genetic ancestry assignment

#### FinnGen

The PCA for population structure has been run using the following approach. First, the following filters were applied: 1) Exclusion of chromosome 23, 2) Exclusion of variants with info score < 0.95, 3) Exclusion of variants with missingness > 0.01, 4) Exclusion of variants with MAF < 0.05, 5) LD pruning with window 500kb, step 50kb, *r*^2^ filter of 0.1.

The imputed genotypes were merged with 1000 genomes phase 3 data into a single data set of 49,451 pruned SNPs, on which the PCs were calculated. An unsupervised Bayesian algorithm (Aberrant) was used to spot outliers in the PCA space and remove them. While this method automatically detected the 1000 genomes samples with non-European and southern European ancestries as outliers, it did not manage to exclude some samples with Western European origins. Since the signal from these samples would have been too small to allow a second round to be performed without detecting substructures of the Finnish population, another approach was used. The FinnGen samples that survived the first round were used to compute another PCA. The European and Finnish 1000 genomes samples were projected onto the space generated by the first 3 PCs. For each sample, the probability of belonging to the EUR/FIN cluster was estimated through using a chi-squared distribution based on the mahalanobis distance to the centroid of each cluster. Samples whose relative probability of being part of the Finnish cluster was > 95% were classified as ethnic Finns and retained in all following analyses for FinnGen (n=309,136).

#### UK Biobank

Principal component analysis and ancestry assignment in UK Biobank followed the same procedure used by the Pan-UK Biobank analysis (<https://pan.ukbb.broadinstitute.org/>) and the procedure is described at <https://pan.ukbb.broadinstitute.org/docs/qc#ancestry-definitions>. We restricted our analysis in UKB to participants of European ancestry (n=426,612).

### Common variant annotation

For variant annotation, we utilized the Variant Effect Predictor^10^ (VEP, <https://www.ensembl.org/info/docs/tools/vep/index.html>). For coding variants, we chose a single most severe consequence and corresponding gene among canonical transcripts. We considered stop_gained, frameshift_variant, splice_donor, splice_acceptor, missense_variant, start_lost, stop_lost, inframe_insertion, and inframe_deletion as protein truncating variants.

### HLA imputation in FinnGen

The HLA imputation is described in detail elsewhere^11^. Briefly, HLA typing on 1150 Finnish samples was performed by the HLA Laboratory of the Finnish Red Cross Blood Service using procedures accredited by the European Federation for Immunogenetics. Allele assignment of the seven classical HLA genes at two-field resolution level (i.e., unique protein sequence level) was performed by polymerase chain reaction (PCR) -based methods. HIBAG^12^ v1.14.0 with 100 classifiers for each of the seven HLA genes was fitted using the training data of 1150 individuals to construct an imputation reference for the Finnish population, which was used to impute the HLA alleles in FinnGen^11^.

### Statistical fine-mapping in FinnGen

Summary statistics for fine-mapping were obtained from standard FinnGen pipeline summary statistics, where mixed model logistic regression using SAIGE^13^ was used to obtain summary statistics for each FinnGen endpoint used to define the 80 diseases (**ST2**). The models used sex and age as precision covariates. Genotyping batch and 10 first genetic principal components were used to control for confounding due to population stratification and batch effects. Using the summary statistics, we fine-mapped all regions with at least one variant having *P*<10^-8^ and extended the regions 1.5 megabases (Mb) upstream and downstream from each lead variant. Overlapping regions were merged and used in SuSiE^14^ fine-mapping, allowing for up to 10 causal variants per region and constructing 95% credible sets for each independent signal. In-sample dosage LD was computed using LDStore2. The FinnGen fine-mapping pipeline is available at <https://github.com/FINNGEN/finemapping-pipeline>.

### Statistical fine-mapping in UK Biobank

The fine-mapping in UK Biobank using SuSiE followed a similar procedure. Regions for fine-mapping were defined by greedily starting with the most significantly associated (highest chi-square) variant, including all genome-wide significant (*P*<10^-8^) variants within a window of 3 Mb centered at the variant, and merging overlapping regions. Summary statistics were obtained using BOLT-LMM^15^ and SAIGE^13^. In-sample dosage LD was estimated using LDStore2. The maximum number of causal variants for each locus was 10. We only considered fine-mapping results for six quantitative risk factor traits (BMI, HbA1c, HDL-C, LDL-C, SBP, cigarettes per day) and considered variants with *P*<10^-12^ to restrict the number of variants to be selected.

### LD clumping common variants

Despite selecting the top posterior inclusion probability common variant out of each credible set for each disease (and the 6 UKB risk factors), the common variants can be in high linkage disequilibrium. This is because we use fine-mapping results for multiple phenotypes to select the initial list of common variants. To select a set of common variants that are independent of each other, we performed LD clumping to list of 2562 variants as follows.

We used *PLINK*^16^ version 1.90b6.24 to clump all common variants using an *r^2^* threshold of 0.2, a 250kb clumping window, and FinnGen release 4 genotypes as the reference panel to remove SNPs in LD with variants having a smaller minimum *P*-value among the 80 hazard ratios for all examined diseases (meta-analysis estimates combining FinnGen and UKB). However, if there was a coding variant among the variants in LD, that was instead kept and others discarded. Additionally, if there was a Finnish-enriched variant (enrichment >5 times) among the variants in LD (but no coding variant), the Finnish-enriched variant was kept. This resulted in 1044 independent common variants that were used in the analyses and we report results for.

### Uncertainty estimation of attributable DALYs

Assuming that there is no uncertainty in the DALY estimates from GBD and the estimated population prevalences of the exposures (e.g., allele frequencies), for a single disease both attributable individual and population DALYs are a deterministic function of the HRs between the exposure and the diseases. Therefore, confidence intervals for the effect of a genetic exposure on attributable DALYs through one disease was estimated using the delta method.

Estimating the *total* attributable DALYs through the 80 examined diseases is less straightforward, as the HRs for different diseases are not independent (e.g., ischemic heart disease and lower extremity peripheral artery disease are comorbid, so risk variants tend to increase risk for both). Bootstrapping was not computationally feasible, so we estimated the uncertainty via resampling the multivariate normal distribution of the log-HR estimates.

Considering a single genetic exposure $e$, let $\boldsymbol{b}_{e}={{(b}_{e,1},\ldots, b_{e,80})}^{T}$ denote the random vector of the log-HRs between exposure $e$ and the $d=1,2,\ldots,80$ diseases, let $\boldsymbol{\beta}_{e}$ denote the estimated vector of Cox model coefficients (log-HRs) for the 80 diseases, let $\boldsymbol{\Sigma}_{e}$ denote the covariance matrix of those coefficients, where the diagonal represents the standard errors of the coefficients. The coefficients follow a multivariate normal distribution:

$$\boldsymbol{b}_{e}\mathcal{\sim N}\left( \boldsymbol{\beta}_{e},\boldsymbol{\Sigma}_{e} \right)$$

We can express the covariance matrix $\boldsymbol{\Sigma}_{e}$ in terms of the diagonal matrix $\boldsymbol{D}_{e}=\mathrm{diag}\left( \boldsymbol{\sigma}_{e} \right)$ that has the standard errors of the coefficients $\boldsymbol{\sigma}_{e}\boldsymbol{=}{{(\sigma}_{e,1},\ldots, \sigma_{e,80})}^{T}$ on the diagonal and the correlation matrix of the coefficients $\boldsymbol{C}$ as:

$$\boldsymbol{\Sigma}_{e}\boldsymbol{=}\boldsymbol{D}_{e}\boldsymbol{C}\boldsymbol{D}_{e}$$

so that

$$\boldsymbol{b}_{e}\mathcal{\sim N}\left( \boldsymbol{\beta}_{e},\boldsymbol{D}_{e}\boldsymbol{C}\boldsymbol{D}_{e} \right)$$

We estimate $\boldsymbol{\beta}_{e}$ and $\boldsymbol{\sigma}_{e}$ from the 80 Cox models for each disease (for common variants we use the meta-analysis estimates from FinnGen and UKB). Let $d=1,2,\ldots,80$ index all the different diseases. We estimated $\boldsymbol{C}_{d\times d}$ by taking all the shrunk log-HRs (assuming they represent null effects) between *all* common variant-disease pairs and calculating the Pearson’s correlation coefficient between the log-HRs of two diseases:

$${\hat{\boldsymbol{C}}}_{i,j}\boldsymbol{=r(}{\hat{\boldsymbol{\beta}}}_{i}\boldsymbol{,}{\hat{\boldsymbol{\beta}}}_{j}\boldsymbol{)}$$

where ${\hat{\boldsymbol{\beta}}}_{i}$ and ${\hat{\boldsymbol{\beta}}}_{j}$ are the Cox model coefficients for disease $i,j=1,2,\ldots,80$ for common variants not shrunk for diseases $i,j$ (at most $1044$). We restricted the correlation estimation to unshrunk variants to to make the coefficients reflect sampling variability, not true effects.

For each genetic exposure $e$ we then resample the vector of log-HRs $B=1,2, \ldots,10 000$ times from the multivariate normal distribution

$$\boldsymbol{b}_{e,B}^{*}\mathcal{\sim N}\left( {\hat{\boldsymbol{\beta}}}_{\boldsymbol{e}}\boldsymbol{,}{\hat{\boldsymbol{D}}}_{e}\hat{\boldsymbol{C}}{\hat{\boldsymbol{D}}}_{e} \right)$$

to emulate the sampling distribution of the vector of log-HRs across all diseases that accounts for dependence in log-HRs between diseases. We then repeat the estimation procedure for individual and population total attributable DALYs as described in main manuscript (**Methods**) for each genetic exposure 10 000 times using the resampled $\boldsymbol{b}_{e,B}^{*}$ to calculate the HRs instead of the maximum partial likelihood estimates from the Cox model (${\hat{\boldsymbol{\beta}}}_{i}$). We then use the 2.5% and 97.5% percentiles of the resampled distribution as estimates of the 95% confidence intervals and calculate the P-values using a normal approximation. In this approach, we do not account for 1) uncertainty in the GBD study estimating population DALYs for each disease, 2) uncertainty in estimating population allele frequencies, and 3) relatedness between individuals.

### Colocalization in FinnGen

As a sensitivity analysis, we performed a colocalization analysis for all fine-mapped regions in FinnGen. For each reported common variant, we checked whether for each pair of diseases the 95% credible sets from SuSiE^14^ fine-mapping results contain the variant. If this was the case, the two diseases colocalize for that variant. We performed this analysis only for the 44 diseases reported in this paper that are defined through a single FinnGen endpoint (**ST2**), as fine-mapping results are not available for the diseases defined through composites of multiple FinnGen endpoints.

### Simulations of underlying effect sizes for examining performance of shrinkage method

We used Hail 0.2 (<https://hail.is/>) to simulate variant-phenotype associations for 80 diseases where the true underlying effects are known. The approach uses genetic data from 361,194 individuals from UK biobank with European ancestry and has the advantage of using realistic variant frequencies and population structure as compared to simulated genetic data. Only 558,240 independent HapMap 3 SNPs were considered in the analysis. Using the *ldscsim.simulate_phenotypes* function we simulated 71 phenotypes based on a spike & slab model with different probability of SNP being causal (*π*). The heritability of the phenotypes was randomly sampled from a uniform distribution ranging from 10 to 60%. The phenotypes were consequently binarized based on the disease prevalence observed in FinGen using the function *ldscsim.binarize.* We considered four *π* values*:* 0.001,0.002,0.005,0.01 meaning that 0.1%, 0.2%,0.5% and 1% of the 558,240 independent variants had a true underlying effect different from 0. The size of this effect is then a function of the heritability of the phenotype.

We then run a GWAS for each of the phenotypes across the 4 different *π* scenarios*.* This allows us to obtain an *observed* effect size from the GWAS and an *expected* true underlying effect. Out of the *observed* effects, consistently with the variant selection process used on the real data, we only included variants that had at least 1 genome-wide significant association (p<5x10^-8^). For this selected group of variants, we applied the same shrinkage procedure as in the main analysis, using the same priors. Our procedure shrinks most of the variant-phenotype associations to 0, while maintaining others unshrunk. Because we know the true underlying effect sizes, that is, which variants have effect size of 0 (null model) and effect sizes different from 0 (alternative model), we can compare how well our procedure shrinks variants from the null model versus does not shrink those from the alternative model. We report the classifier performance in **ST7** and **Extended data Fig. 7**. Overall, our approach results in AUCs of 0.817 to 0.888 for different values of *π*.

### FinnGen ethics statement

Patients and control subjects in FinnGen provided informed consent for biobank research, based on the Finnish Biobank Act. Alternatively, separate research cohorts, collected prior the Finnish Biobank Act came into effect (in September 2013) and start of FinnGen (August 2017), were collected based on study-specific consents and later transferred to the Finnish biobanks after approval by Fimea (Finnish Medicines Agency), the National Supervisory Authority for Welfare and Health. Recruitment protocols followed the biobank protocols approved by Fimea. The Coordinating Ethics Committee of the Hospital District of Helsinki and Uusimaa (HUS) statement number for the FinnGen study is Nr HUS/990/2017.

The FinnGen study is approved by Finnish Institute for Health and Welfare (permit numbers: THL/2031/6.02.00/2017, THL/1101/5.05.00/2017, THL/341/6.02.00/2018, THL/2222/6.02.00/2018, THL/283/6.02.00/2019, THL/1721/5.05.00/2019 and THL/1524/5.05.00/2020), Digital and population data service agency (permit numbers: VRK43431/2017-3, VRK/6909/2018-3, VRK/4415/2019-3), the Social Insurance Institution (permit numbers: KELA 58/522/2017, KELA 131/522/2018, KELA 70/522/2019, KELA 98/522/2019, KELA 134/522/2019, KELA 138/522/2019, KELA 2/522/2020, KELA 16/522/2020), Findata permit numbers THL/2364/14.02/2020, THL/4055/14.06.00/2020,,THL/3433/14.06.00/2020, THL/4432/14.06/2020, THL/5189/14.06/2020, THL/5894/14.06.00/2020, THL/6619/14.06.00/2020, THL/209/14.06.00/2021, THL/688/14.06.00/2021, THL/1284/14.06.00/2021, THL/1965/14.06.00/2021, THL/5546/14.02.00/2020 and Statistics Finland (permit numbers: TK-53-1041-17 and TK/143/07.03.00/2020 (earlier TK-53-90-20)).

The Biobank Access Decisions for FinnGen samples and data utilized in FinnGen Data Freeze 7 include: THL Biobank BB2017_55, BB2017_111, BB2018_19, BB_2018_34, BB_2018_67, BB2018_71, BB2019_7, BB2019_8, BB2019_26, BB2020_1, Finnish Red Cross Blood Service Biobank 7.12.2017, Helsinki Biobank HUS/359/2017, Auria Biobank AB17-5154 and amendment #1 (August 17 2020), Biobank Borealis of Northern Finland_2017_1013, Biobank of Eastern Finland 1186/2018 and amendment 22 § /2020, Finnish Clinical Biobank Tampere MH0004 and amendments (21.02.2020 & 06.10.2020), Central Finland Biobank 1-2017, and Terveystalo Biobank STB 2018001.
